# A Non-parametric Approach to the Construction of Life Tables at the Local Level with Application to Andhra Pradesh, India

**DOI:** 10.1101/2023.03.04.23286753

**Authors:** Aalok Ranjan Chaurasia

## Abstract

In this paper, a non-parametric approach has been proposed for the construction of life table at the local level. The approach is based on the non-parametric modelling of the transition in age-specific probabilities of death at higher levels of administration and the relationship between the geometric mean of the age-specific probabilities of death and the probability of death in the first five years of life. The application of the approach has been demonstrated by constructing life tables for the districts of Andhra Pradesh, India using the data from the 2011 population census and the sample registration system.

## Introduction

A life table is an empirical model that synthesises the mortality experience of the population in a clear and concise manner (Namboodari and Suchindran, 1987). It provides the basis for measuring the longevity in the population. The life expectancy, or the expected number of years of life remaining at a specified age, derived from the life table, is universally the most widely used indicator of population health. A high life expectancy at birth is commonly viewed as a matter of aggrandisement while a low life expectancy at birth reflects the poor quality of the life of the people. Improvement in life expectancy reflects the progress in human well-being. The life expectancy at birth is one of the three constituents of the human development index developed and used by the United Nations to rank countries in terms of human well-being (United Nations, 2022). Life expectancy is also commonly used to assess potential health inequalities across countries, populations, and administrative areas and for evaluating the effectiveness of public health programmes and interventions. Life expectancy and contributes to the development of future healthcare policies and is central for health needs profiling at the local level.

Life tables in India are available for selected states of the country, having a population of more than 20 million, only. They are based on the official sample registration system of the country (Government of India, 2022a). Life tables are not available for the smaller states and Union Territories of the country. Similarly, life tables are also not available for the districts of the country. The non-availability of life tables at the district level is a major impediment to measuring and monitoring population health at the local level and to analyse inter-district health inequalities that appear to be quite pervasive in the country. Constructing life tables and estimating life expectancy at the district level is important in the sense that they may help in identifying the hotspot districts as regards the population health.

Construction of life tables and estimation of life expectancy require population and death counts by age. Although, registration of births and deaths in India is compulsory under the Birth and Death Registration Act of 1969 (Government of India, 1969), yet registration of births and deaths in the country is deficient to provide data necessary for constructing life tables at the district and below district levels. The sample registration system in India was launched in 1967 to provide estimates of key demographic indicators at the state level on an annual basis (Government of India, 2022b) but the system has sample size limitations to provide district level estimates of age-specific death rates required to construct the life tables and estimate life expectancy. In 1992, the Government of India launched the National Family Health Survey Programme (Government of India, 2022c). The first three rounds of the survey provided estimates of key population and health related indicators at the state level only but the fourth and the fifth rounds of the survey have provided estimates of selected population and health related indicators for the districts also. However, because of sample size limitations, the data available through the survey are not adequate to construct life tables and estimate life expectancy at the district level.

The non-availability of district level life tables in India is a major impediment to measuring and monitoring population health at the district level and to analyse the impact of health policies and programme in meeting the health needs of the local people. Monitoring population health at the district level is important as the National Health Mission emphasises district-based decentralised planning for the delivery of health services to meet the health needs of the people (Government of India, 2013). District level life tables is also necessary to highlight inter-district disparities in population health and in identifying hotspot districts which may be given focused attention in planning for the delivery of health care. It is well-known that inter-district disparity in population health in India is quite pervasive and has persisted over time. It has, therefore, been repeatedly argued that reducing inter-district disparities in population health may contribute substantially towards accelerating the pace of improvement in population health in the country. Constructing district life tables is the first step in this direction as it provides the evidence about the magnitude of the problem and then analysing how reduction in the inter-district disparity may contribute to improving the health of the population of the country.

In the absence of the reliable data about population and death counts classified by age and sex at the district level which are necessary for the construction of the life table, indirect approaches have been used to construct life tables at the district level in the country. The first attempt, in this direction, was made by Ponnapalli (2011) who constructed life tables for the districts of the Punjab state of the country using an indirect estimate of life expectancy at birth and model life tables. Kesarwani (2015), on the other hand, has constructed life tables for all districts of the country based on national and state level life tables available through the sample registration system of the country and district level estimates of infant mortality rate derived from the summary birth history data available from the decennial population census. The method followed by Kesarwani (2015) uses the correlation that exists between the infant mortality rate and the life expectancy and birth and the correlation between life expectancy at successive ages. The same approach has also been used by Saikia and Borah (2016) to construct life tables for 23 districts of the Assam state. These studies have first estimated the life expectancy at birth based on the strong correlation between the life expectancy at birth and child mortality, especially, infant mortality, and then constructed life tables by either using the model life tables or using the association between the life expectancy at birth and life expectancy at successive ages. There has been no attempt to first estimate the age-specific probabilities of death at the district level and then to construct life table based on the estimated age-specific probabilities of death and estimating life expectancy.

In this paper, we propose an alternative approach to construct life tables at the district level by first estimating the age-specific probabilities of death in the district and then constructing life table using the estimated age-specific probabilities of death. The age-specific probabilities of death in the district are estimated based on the modelling of the transition in age-specific probabilities of death in the state/Union Territory to which the district belongs. The state level life tables available through the sample registration system have been used for modelling the transition in the age-specific probabilities of death. Since life tables based on the sample registration system are not available for all states/Union Territories of the countries, transition in the age-specific probabilities of death at the national level may be used to estimate age-specific probabilities of death in districts of those states/Union Territories for which life tables are not available from the sample registration system. Once the age-specific probabilities of death are estimated, life tables may be constructed through the standard procedure, for example, by using the MORPAK software package developed by United Nations for mortality analysis (United Nations 2013). The advantage of the approach proposed in this paper is that it allows construction of life tables for sub-groups of the population within the district corresponding to life tables for the population sub-group at the state level.

The paper is organised as follows. The next section of the paper describes the approach proposed to estimate district level age-specific probabilities of death based on the transition in the age-specific probabilities of death in the state to which the district belongs. The third section of the paper applies the proposed approach to construct life tables for the districts of Andhra Pradesh. For each district, life tables have been constructed for the total population and for the eight population sub-groups. The fourth section of the paper analyses the inter-district disparity in population health in Andhra Pradesh. The fifth section analyses within district disparity in population health across the four mutually exclusive and exhaustive population subgroups – rural male, rural female, urban male, and urban female. The last section of the paper discusses the possibility of creating district level mortality database in the country to facilitate the measurement and monitoring of population health at the district level and to identify hotspot districts using the approach of constructing district life tables outlined in the paper.

## The Method

It is well-known that the variation in the age-specific probabilities of death can be captured in terms of the average probability of death across all ages and the age pattern of the probability of death as the risk of death varies by age. If *q*_*yj*_ denotes the probability of death in age *j* in the year *y*, then *q*_*yj*_ can be written as

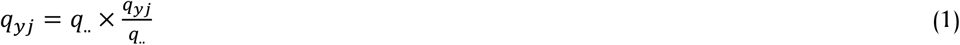

where *q*_.._ is the average probability of death across all years (*y*) and across all ages (*j*).

Let *q*_*y*._ is the average probability of death in the year *y* across all ages *j* and *q*_.*j*_ is the average probability of death in age *j* across all years and

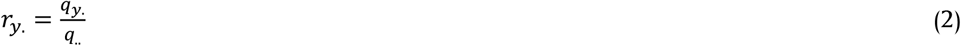

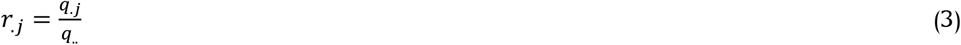

Then, we can write,

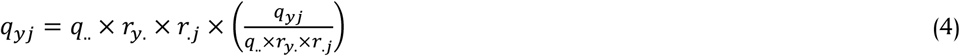

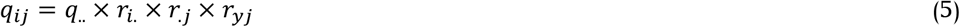

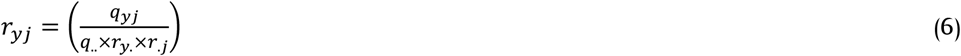

where,

Equation (5) suggests that the probability of death in any age group (*j*) in the year (*y*) can be decomposed into four components: 1) average mortality across all years and all ages (*q*_.._); 2) a year multiplier (*r*_*y*._) which shows that how average mortality in a year is higher or lower than the average mortality across all years; 3) an age multiplier (*r*_.*j*_) which shows how the average mortality in an age is higher or lower than the average mortality of all ages; and 4) the residual term (*r*_*yj*_) which is specific to the year *y* and the age *j*. Equation (5) models the transition in age-specific probabilities of death over years and across age. It may be observed from equation (5) that the values of *q*_.._, and *r*_.*j*_ remain fixed for all values of *y* while *r*_*y*._, and *r*_*yj*_ varies with *y*. This means that for different values of *q*_*y*._, equation (5) yields different values of *q*_*yj*_. The transition in mortality is reflected by both transition in *q*_*y*._ and transition in *r*_*yj*_.

Equation (5) can be fitted by using the data mining technique of polishing by using an appropriate polishing function. The polishing function may be the simple arithmetic mean of age-specific probabilities of death or the geometric mean of the age-specific probabilities of death. Since equation (5) is a multiplicative model, the geometric mean is preferred over the simple arithmetic mean. The geometric mean of age-specific probabilities of death is equal to the simple arithmetic mean of the logarithm of the age-specific probabilities of death.

Equation (5) can be used to estimate age-specific probabilities of death in a district if an estimate of the geometric mean of the age-specific probabilities of death is known. The empirical analysis suggests that there exists near linear relationship between *q*_*y*._ and the probability of death in the first five years of life so that an estimate of the geometric mean of the age-specific probabilities of death can be obtained from the probability of death in the first five years of life assuming that state level linear relationship between the geometric mean of the age-specific probabilities of death and probability of death in the first five years of life holds for the district also. Once the geometric mean of age-specific probabilities of death is estimated, age-specific probabilities of death for the district can be estimated from equation (5).

The probability of death in the first five years of life at the local level can be estimated from the summary birth history data available through the decennial population census using the indirect technique of child mortality estimation (United Nations, 1983; Maultrie et al, 2013). Census-based estimates of the probability of death in the first five years of life, however, refer to period around five-years prior to the date of the population census and, therefore, need to be updated for the most recent date for the purpose of constructing the life table and estimating life expectancy. Chaurasia (2021) has developed a non-parametric approach to combine district level estimates of the probability of death in the first five years of life obtained from the summary birth history data available from the decennial population census and state/Union Territory level estimates of the probability of death in the first five years of life for the most recent date, available through some other source such as the sample registration system to obtain district level estimates of the probability of death in the first five years of life for the most recent date. The approach adopted by Chaurasia (2021) involves empirical modelling of the variation in the probability of death in the first five years of life across mutually exclusive and exhaustive population sub-groups in each district and combining the empirical model with the latest estimates of the probability of death in the first five years of life at the state/Union Territory level available through the sample registration system to obtain district level estimates of the probability of death in the first five years of life for the most recent date. The approach is dynamic in the sense that the new population census may lead to new empirical model of variation in the probability of death in the first five years of life and hence new estimates of the probability of death in the first five years of life for the constituent districts of the state/Union Territory. These estimates of the probability of death in the first five years of life may be used to estimate the geometric mean of age-specific probabilities of death and for the construction of life tables.

The method of constructing the life table at the local (district) level may now be described in terms of the following steps:

1. Using the age-specific probabilities of death for different years available from the sample registration system, construct the mortality transition model for the state in conjunction with equation (5) through the application of the data mining technique of polishing with the geometric mean of the age-specific probabilities of death as the polishing function.

2. Establish the relationship between the geometric mean of the age-specific probabilities of death and the probability of survival in the first five years of life for the state using the age-specific probabilities of death for the state available from the sample registration system.

3. Estimate the probability of death in the first five years of life for each of the constituent districts of the state using the summary birth history data collected at the time of the latest decennial population census by applying the indirect technique of child mortality estimation.

4. Update the estimate of the probability of death in the first five years of life for the districts obtained from the decennial population census corresponding to the most recent estimate of the probability of death in the first five years of life for the state available from the sample registration system or from any other source.

5. Use the updated estimate of the probability of death in the first five years of life for the district to estimate the geometric mean of the age-specific probabilities of death based on the relationship between the geometric mean of the age-specific probabilities of death and the probability of death in the first five years of life in the state.

6. Use the geometric mean of the age-specific probabilities of death for the district to obtain district-specific age-specific probabilities of death in conjunction with the equation (5).

7. Construct the life table for the district from the age-specific probabilities of death using the MORTPAK software package for mortality analysis (United Nations, 2013).

8. In states/Union Territories where life tables from the sample registration system or from any other source are not available, national life tables may be used as the mortality transition model.

## Application to Andhra Pradesh

We demonstrate the application of the approach described in the previous section to construct life tables for the districts of Andhra Pradesh based on the data available through the 2011 population census and sample registration system. Life tables for Andhra Pradesh are available from the sample registration system for the period 1971-75 through 2016-20 and these life tables have been used to develop the mortality transition model for the state in conjunction with equation (5). The resulting mortality transition model for males is presented in table 1 and for females in table 2. It may be seen from table 1 that the average mortality in males of the state (geometric mean) in 1971-75 was almost 88 per cent higher than the average for the period 1971-2020 but, in 2016-20, was around 34 per cent lower than the average for the period 1971-2020. On the other hand, table 2 suggests that average mortality of females in 1971-75 was more than 125 per cent higher than the average for the period 1971-2020 but was almost 39 per cent lower in 2016-20. Similarly, on average, the male probability of death in the age group 0-1 year is almost 47 per cent higher than the average but was only around 12 per cent of the average in the age group 10-14 years. In females, the probability of death in the 0-1year age-group is almost 72 per cent higher than the average but is only around 12 per cent of the average in the age group 10-14 years. Variation from this average in different years is determined by the residuals *r*_*yj*_.

**Table 1:**
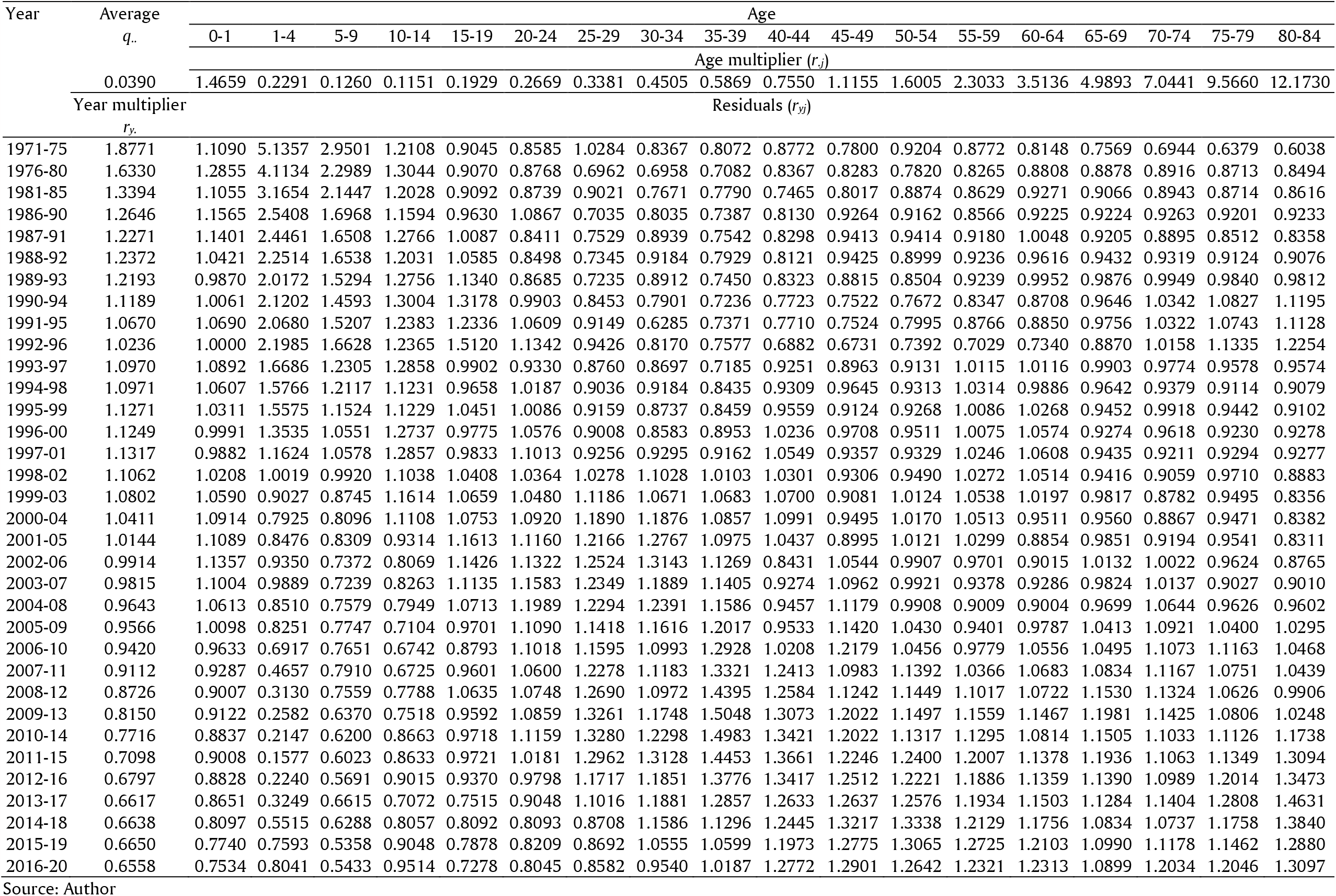
Andhra Pradesh – male mortality transition model, 1971-2020.

**Table 2:** Andhra Pradesh – female mortality transition model, 1971-2020.

| Year | Average<br>$q_{..}$ | Age | | | | | | | | | | | | | | | | | |
| --- | --- | --- | --- | --- | --- | --- | --- | --- | --- | --- | --- | --- | --- | --- | --- | --- | --- | --- | --- |
|  |  | 0-1 | 1-4 | 5-9 | 10-14 | 15-19 | 20-24 | 25-29 | 30-34 | 35-39 | 40-44 | 45-49 | 50-54 | 55-59 | 60-64 | 65-69 | 70-74 | 75-79 | 80-84 |
| | | Age multiplier ($r_{ij}$) | | | | | | | | | | | | | | | | | |
|  | 0.0320 | 1.7173 | 0.3364 | 0.1596 | 0.1187 | 0.2724 | 0.3037 | 0.3221 | 0.3640 | 0.4194 | 0.5686 | 0.8282 | 1.3051 | 1.9915 | 3.2445 | 4.9452 | 7.6647 | 10.6015 | 14.0826 |
| Year multiplier | | Residuals ($r_{ij}$) | | | | | | | | | | | | | | | | | |
| | $r_y$ | | | | | | | | | | | | | | | | | | |
| 1971-75 | 2.2512 | 0.8523 | 4.1153 | 2.3139 | 1.5184 | 1.0931 | 1.1172 | 1.1882 | 1.0813 | 1.1011 | 0.9029 | 0.8806 | 0.8250 | 0.7463 | 0.7960 | 0.6745 | 0.5990 | 0.5641 | 0.5288 |
| 1976-80 | 1.7998 | 1.0465 | 3.8132 | 2.2221 | 1.2827 | 0.9819 | 1.0386 | 1.0961 | 1.0306 | 0.9512 | 0.8479 | 0.8059 | 0.7987 | 0.7793 | 0.8154 | 0.7481 | 0.6927 | 0.6794 | 0.6571 |
| 1981-85 | 1.5046 | 0.9250 | 2.6123 | 1.6574 | 1.3785 | 1.0349 | 1.0199 | 1.0413 | 1.0003 | 0.8800 | 0.8965 | 0.9447 | 0.8224 | 0.7184 | 0.7991 | 0.8036 | 0.8046 | 0.8465 | 0.8556 |
| 1986-90 | 1.3750 | 0.9883 | 2.5061 | 1.4329 | 1.2995 | 1.0297 | 1.1370 | 0.9261 | 1.0233 | 0.8935 | 0.8750 | 0.7752 | 0.8333 | 0.7358 | 0.8734 | 0.8614 | 0.8596 | 0.8981 | 0.9054 |
| 1987-91 | 1.3192 | 0.9659 | 2.3427 | 1.4192 | 1.2945 | 1.0471 | 0.9774 | 0.9549 | 0.9382 | 0.9477 | 0.9057 | 0.7871 | 0.8496 | 0.8000 | 0.8922 | 0.8835 | 0.8730 | 0.9075 | 0.9147 |
| 1988-92 | 1.3058 | 0.9297 | 2.2656 | 1.2341 | 1.2775 | 1.0798 | 0.9914 | 0.9573 | 0.9215 | 0.9129 | 0.8902 | 0.7741 | 0.7943 | 0.8331 | 0.8693 | 0.9259 | 0.9419 | 0.9995 | 1.0111 |
| 1989-93 | 1.3424 | 0.8447 | 1.8062 | 1.1785 | 1.3801 | 1.1137 | 1.0273 | 0.9949 | 0.9694 | 0.9180 | 0.8700 | 0.7898 | 0.7922 | 0.8343 | 0.8083 | 0.9382 | 0.9897 | 1.0745 | 1.0862 |
| 1990-94 | 1.4222 | 0.8880 | 1.3145 | 1.2076 | 1.1915 | 0.7872 | 0.7741 | 0.7909 | 0.9597 | 1.0548 | 1.1782 | 1.1689 | 1.0328 | 1.0907 | 1.1180 | 1.0067 | 0.9209 | 0.8912 | 0.8517 |
| 1991-95 | 1.4218 | 0.9061 | 1.2409 | 1.0921 | 1.1270 | 0.9635 | 0.8352 | 0.8929 | 1.0876 | 1.0394 | 1.0768 | 1.0981 | 1.0181 | 1.0834 | 1.0874 | 0.9837 | 0.8949 | 0.8636 | 0.8251 |
| 1992-96 | 1.3045 | 0.8864 | 1.3582 | 1.1903 | 1.1475 | 0.9199 | 0.8716 | 0.9322 | 1.0529 | 1.0625 | 1.0420 | 1.0404 | 0.9974 | 0.9797 | 0.9295 | 0.9450 | 0.9079 | 0.9291 | 0.9267 |
| 1993-97 | 1.2053 | 0.8920 | 1.3187 | 1.2963 | 1.2857 | 1.1413 | 1.0322 | 0.9815 | 0.8785 | 0.8974 | 0.9178 | 0.9439 | 1.0245 | 0.8322 | 0.8804 | 0.9249 | 0.9288 | 0.9884 | 1.0134 |
| 1994-98 | 1.1712 | 0.9516 | 1.3325 | 1.3007 | 1.1454 | 0.7961 | 0.9321 | 0.9487 | 1.0421 | 1.0707 | 0.9399 | 1.0031 | 1.0679 | 0.8446 | 0.9201 | 0.9385 | 0.9311 | 0.9817 | 1.0040 |
| 1995-99 | 1.1472 | 1.0054 | 1.2963 | 1.3705 | 1.0659 | 1.1501 | 0.9157 | 0.9846 | 1.0676 | 0.9428 | 0.9149 | 1.0095 | 1.0971 | 0.8949 | 0.9221 | 0.8051 | 0.8917 | 0.8787 | 0.9540 |
| 1996-00 | 1.1249 | 1.0066 | 1.1376 | 1.2844 | 1.0988 | 1.1320 | 0.9650 | 1.0171 | 1.0735 | 0.9416 | 0.8871 | 1.0265 | 1.0835 | 0.9048 | 0.9264 | 0.8460 | 0.9251 | 0.8728 | 0.9753 |
| 1997-01 | 1.0619 | 1.0765 | 0.9669 | 1.1132 | 1.1639 | 1.1624 | 1.0173 | 1.0683 | 1.0498 | 0.9111 | 0.8812 | 0.9572 | 1.1006 | 0.9585 | 0.9264 | 0.8961 | 0.9594 | 0.8776 | 0.9867 |
| 1998-02 | 1.0262 | 1.1131 | 0.9752 | 1.1902 | 1.0247 | 1.1033 | 1.0437 | 1.0827 | 1.0486 | 0.9246 | 0.9091 | 0.9386 | 1.0296 | 0.9531 | 0.9247 | 0.9170 | 0.9761 | 0.9098 | 0.9911 |
| 1999-03 | 0.9666 | 1.1842 | 0.9371 | 1.1723 | 1.0061 | 1.0941 | 1.0921 | 1.1143 | 0.9728 | 0.9322 | 0.8787 | 0.8742 | 1.0025 | 0.9950 | 0.9124 | 0.9019 | 1.0453 | 0.9697 | 0.9901 |
| 2000-04 | 0.9198 | 1.2164 | 0.9363 | 1.0849 | 0.9856 | 1.0886 | 1.2193 | 1.1077 | 0.9531 | 0.9278 | 0.8995 | 0.8862 | 0.9204 | 1.0250 | 0.9030 | 0.9401 | 1.0535 | 0.9550 | 0.9832 |
| 2001-05 | 0.9076 | 1.2343 | 0.9612 | 1.0131 | 1.0715 | 1.0780 | 1.2414 | 1.2029 | 0.9376 | 0.9846 | 0.8965 | 0.8207 | 0.8855 | 1.0241 | 0.8868 | 0.9282 | 1.0372 | 0.9643 | 0.9511 |
| 2002-06 | 0.8752 | 1.2580 | 0.9723 | 0.9386 | 0.9907 | 1.1559 | 1.2120 | 1.0987 | 0.9192 | 1.0501 | 0.7373 | 1.0414 | 0.8728 | 1.0324 | 0.8785 | 0.9567 | 1.0601 | 0.9687 | 0.9909 |
| 2003-07 | 0.8574 | 1.2571 | 0.9492 | 0.7752 | 0.9683 | 1.2402 | 1.2012 | 1.0875 | 0.9824 | 1.0293 | 0.7019 | 0.9983 | 0.9561 | 1.1057 | 0.9407 | 0.9594 | 1.0714 | 0.9284 | 1.0248 |
| 2004-08 | 0.8553 | 1.2070 | 0.9080 | 0.7427 | 0.9553 | 1.2634 | 1.1620 | 1.0902 | 1.0552 | 0.9716 | 0.8246 | 1.0008 | 0.9504 | 1.0225 | 0.9627 | 1.0371 | 1.0110 | 0.9159 | 1.0571 |
| 2005-09 | 0.8560 | 1.1591 | 0.8475 | 0.6642 | 0.9975 | 1.1952 | 1.0660 | 1.0950 | 1.0141 | 0.9751 | 0.8303 | 0.9933 | 0.9644 | 0.9981 | 1.0162 | 1.1258 | 1.0365 | 1.0753 | 1.1003 |
| 2006-10 | 0.8289 | 1.1232 | 0.7103 | 0.5558 | 0.8744 | 1.1872 | 1.1133 | 1.0851 | 1.0318 | 0.9755 | 0.8774 | 1.0664 | 1.0379 | 1.0470 | 1.0878 | 1.2048 | 1.0712 | 1.1259 | 1.1118 |
| 2007-11 | 0.8251 | 1.0807 | 0.6290 | 0.7128 | 0.8943 | 1.1579 | 1.1496 | 1.1018 | 1.0365 | 0.9176 | 1.0354 | 0.8689 | 1.0414 | 1.0301 | 1.1290 | 1.1631 | 1.0737 | 1.1259 | 1.0799 |
| 2008-12 | 0.7894 | 1.0640 | 0.5219 | 0.7699 | 0.8179 | 1.0517 | 1.1559 | 1.1578 | 0.9810 | 1.0111 | 1.1512 | 0.9455 | 0.9768 | 1.0486 | 1.1578 | 1.1996 | 1.1136 | 1.1485 | 1.0345 |
| 2009-13 | 0.7393 | 1.0758 | 0.4265 | 0.7425 | 0.7486 | 0.9910 | 1.1521 | 1.1246 | 0.9673 | 1.0645 | 1.1705 | 1.0326 | 1.0349 | 1.1287 | 1.2242 | 1.2338 | 1.1967 | 1.1688 | 0.9944 |
| 2010-14 | 0.7230 | 1.0241 | 0.3898 | 0.7592 | 0.7654 | 0.9497 | 1.0996 | 1.0169 | 1.0354 | 1.1648 | 1.3315 | 1.1117 | 1.0741 | 1.1384 | 1.1688 | 1.1412 | 1.1351 | 1.1542 | 1.0642 |
| 2011-15 | 0.6851 | 1.0151 | 0.3638 | 0.7583 | 0.7885 | 0.8531 | 0.9515 | 0.9399 | 0.9622 | 1.1802 | 1.4011 | 1.1705 | 1.1506 | 1.2188 | 1.2209 | 1.1501 | 1.1677 | 1.1885 | 1.1662 |
| 2012-16 | 0.6645 | 0.9568 | 0.4185 | 0.7376 | 0.5950 | 0.7932 | 0.8649 | 0.8961 | 0.9663 | 1.2507 | 1.5308 | 1.2211 | 1.1969 | 1.2532 | 1.2410 | 1.2088 | 1.2082 | 1.2277 | 1.2278 |
| 2013-17 | 0.6599 | 0.9007 | 0.4045 | 0.6684 | 0.6590 | 0.8161 | 0.8256 | 0.8434 | 0.9730 | 1.1643 | 1.5047 | 1.2605 | 1.1998 | 1.2778 | 1.2415 | 1.2212 | 1.2528 | 1.3216 | 1.3262 |
| 2014-18 | 0.6526 | 0.8072 | 0.4375 | 0.6158 | 0.7269 | 0.7109 | 0.7323 | 0.8320 | 1.0102 | 1.1099 | 1.4423 | 1.4441 | 1.3002 | 1.3900 | 1.2503 | 1.2745 | 1.1881 | 1.3533 | 1.3504 |
| 2015-19 | 0.6415 | 0.7625 | 0.5379 | 0.6265 | 0.7190 | 0.7053 | 0.7771 | 0.7783 | 0.9674 | 1.0012 | 1.2941 | 1.3544 | 1.2906 | 1.4085 | 1.3332 | 1.3656 | 1.2819 | 1.3914 | 1.2662 |
| 2016-20 | 0.6122 | 0.7308 | 0.5575 | 0.5604 | 0.6888 | 0.6265 | 0.6796 | 0.7996 | 1.0558 | 0.9772 | 1.2984 | 1.4586 | 1.3179 | 1.4384 | 1.3947 | 1.3913 | 1.3455 | 1.4697 | 1.3143 |

Source: Author

On the other hand, the regression of *r*_*y*._ on the probability of death in the first five years of life (*q*_*y5*_) returned the following relationship for males

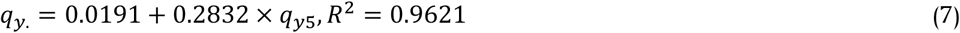

and

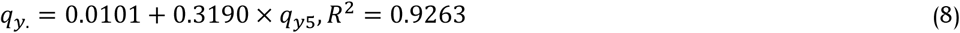

for females and the regression coefficients were highly statistically significant and in the expected direction. Equations (7) and (8) suggest that the geometric mean of age-specific probabilities of death can be estimated fairly accurately from the probability of death in the first five years of life.

Estimates of the probability of death in the first five years of life for the period 2019-20 have been obtained for males and females for the districts of Andhra Pradesh as they existed at the time of 2011 population census by Chaurasia (2022). These estimates are presented in table 3 along with the implied geometric mean of the age-specific probabilities of death in conjunction with equations (7) and (8). The mortality, as reflected by the geometric mean of the age-specific probabilities of death, is estimated to be the lowest in district Sri Potti Sriramulu Nellore but the highest in district Vizainagarm of the state. The table also shows that mortality varies widely across the districts of the state.

**Table 3:**
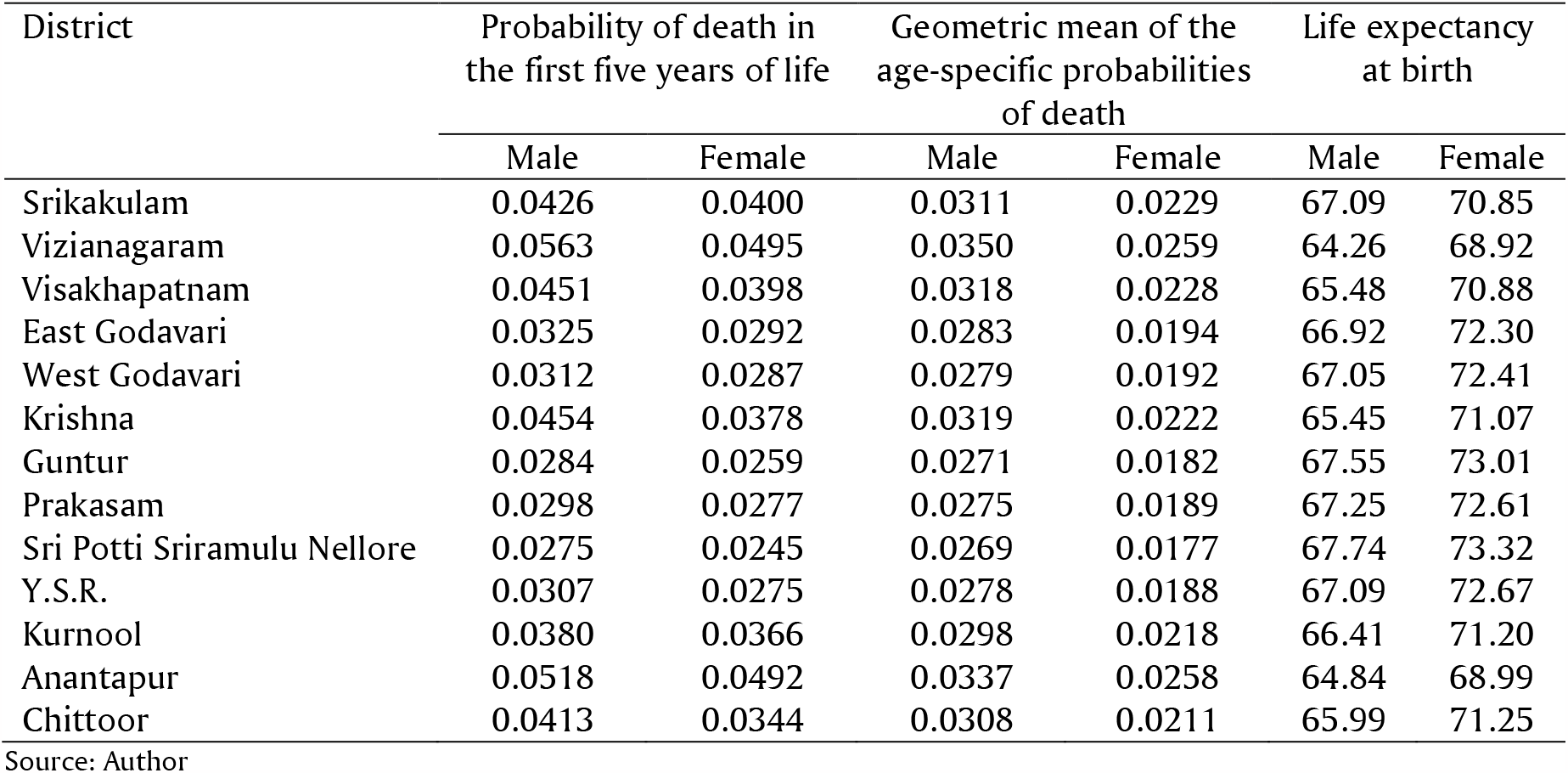
The probability of death in the first five years of life, geometric mean of the age-specific probabilities of death and life expectancy at birth in districts of Andhra Pradesh.

| District | Probability of death in the first five years of life |  | Geometric mean of the age-specific probabilities of death |  | Life expectancy at birth |  |
| --- | --- | --- | --- | --- | --- | --- |
|  | Male | Female | Male | Female | Male | Female |
| Srikakulam | 0.0426 | 0.0400 | 0.0311 | 0.0229 | 67.09 | 70.85 |
| Vizianagaram | 0.0563 | 0.0495 | 0.0350 | 0.0259 | 64.26 | 68.92 |
| Visakhapatnam | 0.0451 | 0.0398 | 0.0318 | 0.0228 | 65.48 | 70.88 |
| East Godavari | 0.0325 | 0.0292 | 0.0283 | 0.0194 | 66.92 | 72.30 |
| West Godavari | 0.0312 | 0.0287 | 0.0279 | 0.0192 | 67.05 | 72.41 |
| Krishna | 0.0454 | 0.0378 | 0.0319 | 0.0222 | 65.45 | 71.07 |
| Guntur | 0.0284 | 0.0259 | 0.0271 | 0.0182 | 67.55 | 73.01 |
| Prakasam | 0.0298 | 0.0277 | 0.0275 | 0.0189 | 67.25 | 72.61 |
| Sri Potti Sriramulu Nellore | 0.0275 | 0.0245 | 0.0269 | 0.0177 | 67.74 | 73.32 |
| Y.S.R. | 0.0307 | 0.0275 | 0.0278 | 0.0188 | 67.09 | 72.67 |
| Kurnool | 0.0380 | 0.0366 | 0.0298 | 0.0218 | 66.41 | 71.20 |
| Anantapur | 0.0518 | 0.0492 | 0.0337 | 0.0258 | 64.84 | 68.99 |
| Chittoor | 0.0413 | 0.0344 | 0.0308 | 0.0211 | 65.99 | 71.25 |

Source: Author

The age-specific probabilities of death for males and females for different districts of the state, obtained using the method described above, are presented in table 4. These age-specific probabilities of death are derived assuming that the mortality transition model of the state presented in tables 1 and 2 holds for the district also so that given the estimated value of the geometric mean of the age-specific probabilities of death in the district, age-specific probabilities of death in the district can be generated. If the geometric mean of the probabilities of death in a district is higher than the geometric mean of the age-specific probabilities of death of the another district, then this implies that the first district is at an earlier stage of mortality transition as compared to the mortality transition in the second district so that the age pattern of mortality in the first district is different from the age pattern of mortality in the second district. This implies that the age specific probabilities of death in the district are shaped by not only the geometric mean of the age-specific probabilities of death estimated on the basis of the estimated probability of death in the first five years of life in the district but also the age pattern of mortality which is different for different value of the geometric mean of age-specific probabilities of death as revealed through the mortality transition model of the state.

**Table 4:** Estimated age-specific probabilities of death in districts of Andhra Pradesh.

| District | Age |  |  |  |  |  |  |  |  |  |  |  |  |  |  |  |  |  |
| --- | --- | --- | --- | --- | --- | --- | --- | --- | --- | --- | --- | --- | --- | --- | --- | --- | --- | --- |
|  | 0-1 | 1-4 | 5-9 | 10-14 | 15-19 | 20-24 | 25-29 | 30-34 | 35-39 | 40-44 | 45-49 | 50-54 | 55-59 | 60-64 | 65-69 | 70-74 | 75-79 | 80-84 |
| Male |  |  |  |  |  |  |  |  |  |  |  |  |  |  |  |  |  |  |
| Srikakulam | 0.04110 | 0.00172 | 0.00247 | 0.00285 | 0.00579 | 0.00912 | 0.01396 | 0.01677 | 0.02744 | 0.03104 | 0.04173 | 0.05691 | 0.08211 | 0.12258 | 0.18313 | 0.24707 | 0.32540 | 0.41032 |
| Vizianagaram | 0.04714 | 0.00330 | 0.00343 | 0.00286 | 0.00673 | 0.00995 | 0.01470 | 0.01751 | 0.02815 | 0.03296 | 0.04323 | 0.06392 | 0.08542 | 0.13154 | 0.19348 | 0.27668 | 0.35845 | 0.43669 |
| Visakhapatnam | 0.04257 | 0.00189 | 0.00257 | 0.00276 | 0.00591 | 0.00922 | 0.01426 | 0.01682 | 0.02809 | 0.03140 | 0.04263 | 0.05858 | 0.08467 | 0.12807 | 0.19016 | 0.25619 | 0.32899 | 0.39687 |
| East Godavari | 0.03716 | 0.00111 | 0.00216 | 0.00281 | 0.00530 | 0.00786 | 0.01246 | 0.01646 | 0.02420 | 0.02903 | 0.03844 | 0.05490 | 0.07705 | 0.11164 | 0.16687 | 0.22016 | 0.30544 | 0.43918 |
| West Godavari | 0.03678 | 0.00104 | 0.00212 | 0.00277 | 0.00523 | 0.00765 | 0.01226 | 0.01641 | 0.02375 | 0.02874 | 0.03805 | 0.05493 | 0.07675 | 0.11104 | 0.16562 | 0.21741 | 0.30241 | 0.44048 |
| Krishna | 0.04266 | 0.00192 | 0.00259 | 0.00277 | 0.00595 | 0.00925 | 0.01428 | 0.01682 | 0.02812 | 0.03144 | 0.04264 | 0.05874 | 0.08475 | 0.12810 | 0.19040 | 0.25681 | 0.32969 | 0.39745 |
| Guntur | 0.03547 | 0.00118 | 0.00200 | 0.00275 | 0.00500 | 0.00724 | 0.01134 | 0.01529 | 0.02249 | 0.02773 | 0.03743 | 0.05344 | 0.07464 | 0.10832 | 0.15796 | 0.21065 | 0.30265 | 0.43822 |
| Prakasam | 0.03624 | 0.00105 | 0.00207 | 0.00275 | 0.00513 | 0.00744 | 0.01190 | 0.01606 | 0.02319 | 0.02831 | 0.03770 | 0.05450 | 0.07600 | 0.10998 | 0.16284 | 0.21424 | 0.30116 | 0.44037 |
| Sri Potti Sriramulu Nellore | 0.03497 | 0.00125 | 0.00196 | 0.00275 | 0.00491 | 0.00711 | 0.01098 | 0.01481 | 0.02204 | 0.02736 | 0.03725 | 0.05277 | 0.07376 | 0.10725 | 0.15485 | 0.20835 | 0.30351 | 0.43678 |
| Y.S.R. | 0.03665 | 0.00102 | 0.00211 | 0.00276 | 0.00521 | 0.00757 | 0.01218 | 0.01639 | 0.02359 | 0.02863 | 0.03791 | 0.05493 | 0.07663 | 0.11081 | 0.16517 | 0.21641 | 0.30130 | 0.44091 |
| Kurnool | 0.03873 | 0.00143 | 0.00232 | 0.00297 | 0.00559 | 0.00880 | 0.01336 | 0.01665 | 0.02614 | 0.03029 | 0.04009 | 0.05459 | 0.07813 | 0.11399 | 0.17193 | 0.23193 | 0.31820 | 0.43151 |
| Anantapur | 0.04461 | 0.00236 | 0.00315 | 0.00301 | 0.00683 | 0.00969 | 0.01456 | 0.01683 | 0.02867 | 0.03221 | 0.04269 | 0.06184 | 0.08615 | 0.12824 | 0.19505 | 0.26939 | 0.34365 | 0.40860 |
| Chittoor | 0.04036 | 0.00163 | 0.00243 | 0.00291 | 0.00574 | 0.00906 | 0.01380 | 0.01674 | 0.02710 | 0.03085 | 0.04125 | 0.05607 | 0.08077 | 0.11969 | 0.17948 | 0.24244 | 0.32365 | 0.41739 |
| Female |  |  |  |  |  |  |  |  |  |  |  |  |  |  |  |  |  |  |
| Srikakulam | 0.04019 | 0.00296 | 0.00277 | 0.00209 | 0.00581 | 0.00745 | 0.00739 | 0.00851 | 0.01121 | 0.01750 | 0.02128 | 0.03251 | 0.05257 | 0.08752 | 0.12936 | 0.20019 | 0.28160 | 0.34912 |
| Vizianagaram | 0.04780 | 0.00511 | 0.00304 | 0.00266 | 0.00787 | 0.00907 | 0.00939 | 0.00957 | 0.01038 | 0.01595 | 0.01931 | 0.03435 | 0.05355 | 0.09591 | 0.15095 | 0.21648 | 0.31186 | 0.38749 |
| Visakhapatnam | 0.04001 | 0.00294 | 0.00276 | 0.00209 | 0.00575 | 0.00735 | 0.00732 | 0.00844 | 0.01118 | 0.01750 | 0.02128 | 0.03255 | 0.05263 | 0.08746 | 0.12895 | 0.19985 | 0.28114 | 0.35012 |
| East Godavari | 0.02432 | 0.00362 | 0.00173 | 0.00158 | 0.00330 | 0.00400 | 0.00500 | 0.00748 | 0.00797 | 0.01439 | 0.02357 | 0.03353 | 0.05588 | 0.08826 | 0.13419 | 0.20109 | 0.30399 | 0.36084 |
| West Godavari | 0.02399 | 0.00355 | 0.00169 | 0.00156 | 0.00324 | 0.00394 | 0.00494 | 0.00742 | 0.00790 | 0.01431 | 0.02346 | 0.03335 | 0.05562 | 0.08783 | 0.13353 | 0.20004 | 0.30265 | 0.35887 |
| Krishna | 0.03876 | 0.00276 | 0.00269 | 0.00206 | 0.00529 | 0.00665 | 0.00685 | 0.00791 | 0.01095 | 0.01747 | 0.02125 | 0.03280 | 0.05302 | 0.08701 | 0.12596 | 0.19728 | 0.27766 | 0.35688 |
| Guntur | 0.02225 | 0.00318 | 0.00152 | 0.00144 | 0.00296 | 0.00362 | 0.00462 | 0.00710 | 0.00752 | 0.01387 | 0.02287 | 0.03235 | 0.05419 | 0.08544 | 0.12988 | 0.19424 | 0.29518 | 0.34804 |
| Prakasam | 0.02339 | 0.00342 | 0.00163 | 0.00152 | 0.00314 | 0.00383 | 0.00483 | 0.00731 | 0.00777 | 0.01416 | 0.02327 | 0.03301 | 0.05515 | 0.08703 | 0.13231 | 0.19809 | 0.30016 | 0.35522 |
| Sri Potti Sriramulu Nellore | 0.02140 | 0.00301 | 0.00144 | 0.00138 | 0.00282 | 0.00347 | 0.00446 | 0.00694 | 0.00732 | 0.01364 | 0.02256 | 0.03182 | 0.05343 | 0.08417 | 0.12795 | 0.19119 | 0.29116 | 0.34236 |
| Y.S.R. | 0.02322 | 0.00339 | 0.00162 | 0.00151 | 0.00312 | 0.00380 | 0.00480 | 0.00728 | 0.00773 | 0.01412 | 0.02321 | 0.03292 | 0.05501 | 0.08680 | 0.13196 | 0.19754 | 0.29945 | 0.35418 |
| Kurnool | 0.03769 | 0.00273 | 0.00263 | 0.00197 | 0.00501 | 0.00622 | 0.00655 | 0.00764 | 0.01089 | 0.01761 | 0.02127 | 0.03294 | 0.05314 | 0.08658 | 0.12495 | 0.19615 | 0.27608 | 0.36089 |
| Anantapur | 0.04756 | 0.00501 | 0.00305 | 0.00263 | 0.00778 | 0.00904 | 0.00939 | 0.00949 | 0.01043 | 0.01603 | 0.01938 | 0.03403 | 0.05343 | 0.09576 | 0.15078 | 0.21634 | 0.31121 | 0.38461 |
| Chittoor | 0.03271 | 0.00287 | 0.00226 | 0.00164 | 0.00469 | 0.00530 | 0.00574 | 0.00747 | 0.01033 | 0.01807 | 0.02200 | 0.03304 | 0.05366 | 0.08499 | 0.12738 | 0.20235 | 0.29489 | 0.39301 |

Source: Author

Finally, using the age-specific probabilities of death presented in table 4, life tables have been constructed separately for male and female population of each district of the state as it existed at the 2011 population census using the MORTPAK software package of mortality analysis (United Nations, 2013). The life tables so constructed for the districts of the state are presented in the appendix tables appended to this paper while the life expectancy at birth for males and females in each district derived from the constructed life tables are presented in table 3.

It may be seen from table 3 that, in the male population, the life expectancy at birth varies from almost 68 years in district Sri Potti Sriramulu Nellore to just around 64 years in district Vizianagarm. In the female population also, the life expectancy at birth varies from almost 73 years in district Sri Potti Sriramulu Nellore to just around 69 years in district Vizianagarm. In Anantapur districts of the state also, the female life expectancy at birth is estimated to be just around 69 years.

The male-female gap in the life expectancy at birth also varies across the districts of the state. In all districts of the state, the life expectancy at birth for the female population is higher than the life expectancy at birth in the male population. In majority of the districts, the female-male gap in the life expectancy at birth is more than 5 years with the highest gap in district Krishna. There are only four districts in the state where the female-male gap in the life expectancy at birth is less than 5 years. On the other hand, Srikakulam is the only district in the state where the female-male gap in the life expectancy at birth is less the 4 years.

It is clear from table 3 and from the life tables constructed for each district of the state that the mortality profile of different districts of the state is essentially different. This implies that population health varies across the districts of the state. It appears that there are district-specific factors that impact the population health of the district. The present analysis emphasises that these district-specific factors should be identified and district level planning and programming for the delivery of health care services should take into consideration these district-specific factors.

## Conclusions

In this paper, I have proposed a non-parametric approach of constructing life tables at the local level in the absence of reliable data on population and deaths classified by age and sex of the population. The method proposed here is particularly suited to situations where the civil registration system is deficient to provide reliable data about population and death counts and there is no other source of data about the population and death classified by age and sex so as to calculate age-specific death rates at the district level which are necessary to construct the life table. In recent years many countries, including India, have carried out sample survey of households to generate data related to population health. However, construction of life tables is not possible from the data available from these surveys because the size of the sample of households surveyed at the local level is too small to provide reliable estimates of age-specific probabilities of death at the local level. In such situations, the only option available is to construct life tables through indirect approaches. The method proposed in this paper is an attempt in this direction.

The method of life table construction at the local level proposed in this paper is a synthetic method. It is based on modelling the transition in mortality and establishing the relationship between the geometric mean of age-specific probabilities of death and the probability of death at the upper level of administrative at which reliable information about mortality is available. The method assumes that the relationship between the geometric mean of the age-specific probabilities of death and the probability of death in the first five years of life that holds at the upper level of administrative also holds for the local level. An advantage of the method proposed here is that it can also be used to construct life tables and estimate life expectancy at birth for different population sub-groups at the local level so as to analyse and highlight local level disparities in population health.

Application of the proposed method to Andhra Pradesh state of India suggests revealing differences in population health across the districts of the state. The life expectancy at birth varies not only across the districts of the state but also across males and females in each district. The gender gap in population health, as revealed through the female-male difference in the life expectancy at birth also varies across the districts of the state. These findings can contribute significantly towards planning and programming for the delivery of health care services at the district level to meet the health needs of the people of the district.

The method of constructing life tables at local level proposed in this paper can be used to develop local level life table database to facilitate research in mortality at the local level and to facilitate evidence-based planning and programming for the delivery of health care services at the local level. Such a database can also help in measuring and monitoring population health at the local level and in highlighting local level disparities in population health.

## Data Availability

All data produced in the present study are available upon reasonable request to the authors

---

Life Tables: District Srikakulum

| Age | m(x,n) | q(x,n) | l(x) | d(x,n) | L(x,n) | T(x) | e(x) | a(x,n) |
| --- | --- | --- | --- | --- | --- | --- | --- | --- |
| Male |  |  |  |  |  |  |  |  |
| 0 | 0.03781 | 0.03665 | 100000 | 3665 | 96921 | 6709302 | 67.09 | 0.16 |
| 1 | 0.00025 | 0.00102 | 96335 | 98 | 385093 | 6612381 | 68.64 | 1.46 |
| 5 | 0.00042 | 0.00211 | 96237 | 203 | 480679 | 6227288 | 64.71 | 2.50 |
| 10 | 0.00055 | 0.00276 | 96034 | 265 | 479508 | 5746609 | 59.84 | 2.50 |
| 15 | 0.00104 | 0.00521 | 95769 | 499 | 477703 | 5267101 | 55.00 | 2.71 |
| 20 | 0.00152 | 0.00757 | 95270 | 722 | 474674 | 4789398 | 50.27 | 2.67 |
| 25 | 0.00245 | 0.01218 | 94549 | 1152 | 470045 | 4314724 | 45.63 | 2.66 |
| 30 | 0.00330 | 0.01639 | 93397 | 1530 | 463361 | 3844679 | 41.16 | 2.63 |
| 35 | 0.00477 | 0.02359 | 91867 | 2167 | 454148 | 3381317 | 36.81 | 2.61 |
| 40 | 0.00581 | 0.02863 | 89699 | 2568 | 442303 | 2927170 | 32.63 | 2.59 |
| 45 | 0.00772 | 0.03791 | 87131 | 3303 | 427802 | 2484867 | 28.52 | 2.62 |
| 50 | 0.01128 | 0.05493 | 83828 | 4605 | 408214 | 2057065 | 24.54 | 2.63 |
| 55 | 0.01591 | 0.07663 | 79223 | 6071 | 381661 | 1648851 | 20.81 | 2.62 |
| 60 | 0.02340 | 0.11081 | 73152 | 8106 | 346476 | 1267190 | 17.32 | 2.62 |
| 65 | 0.03591 | 0.16517 | 65046 | 10744 | 299197 | 920714 | 14.15 | 2.58 |
| 70 | 0.04844 | 0.21641 | 54303 | 11752 | 242610 | 621516 | 11.45 | 2.54 |
| 75 | 0.07080 | 0.30130 | 42551 | 12821 | 181095 | 378906 | 8.90 | 2.53 |
| 80 | 0.11388 | 0.44091 | 29730 | 13108 | 115105 | 197811 | 6.65 | 2.44 |
| 85 | 0.20098 | ... | 16622 | 16622 | 82706 | 82706 | 4.98 | 4.98 |
| Female |  |  |  |  |  |  |  |  |
| 0 | 0.04158 | 0.04019 | 100000 | 4019 | 96667 | 7085281 | 70.85 | 0.17 |
| 1 | 0.00074 | 0.00296 | 95981 | 284 | 383202 | 6988614 | 72.81 | 1.46 |
| 5 | 0.00055 | 0.00277 | 95697 | 265 | 477822 | 6605412 | 69.02 | 2.50 |
| 10 | 0.00042 | 0.00209 | 95432 | 199 | 476660 | 6127590 | 64.21 | 2.50 |
| 15 | 0.00117 | 0.00581 | 95232 | 553 | 474924 | 5650930 | 59.34 | 2.76 |
| 20 | 0.00150 | 0.00745 | 94679 | 705 | 471665 | 5176006 | 54.67 | 2.55 |
| 25 | 0.00148 | 0.00739 | 93974 | 694 | 468150 | 4704341 | 50.06 | 2.52 |
| 30 | 0.00171 | 0.00851 | 93279 | 794 | 464478 | 4236191 | 45.41 | 2.58 |
| 35 | 0.00225 | 0.01121 | 92485 | 1037 | 459987 | 3771713 | 40.78 | 2.65 |
| 40 | 0.00353 | 0.01750 | 91449 | 1600 | 453446 | 3311726 | 36.21 | 2.63 |
| 45 | 0.00430 | 0.02128 | 89848 | 1912 | 444694 | 2858280 | 31.81 | 2.62 |
| 50 | 0.00660 | 0.03251 | 87936 | 2859 | 433043 | 2413586 | 27.45 | 2.68 |
| 55 | 0.01078 | 0.05257 | 85078 | 4473 | 415054 | 1980543 | 23.28 | 2.69 |
| 60 | 0.01825 | 0.08752 | 80605 | 7055 | 386501 | 1565490 | 19.42 | 2.66 |
| 65 | 0.02756 | 0.12936 | 73550 | 9514 | 345178 | 1178989 | 16.03 | 2.63 |
| 70 | 0.04432 | 0.20019 | 64036 | 12819 | 289262 | 833811 | 13.02 | 2.59 |
| 75 | 0.06555 | 0.28160 | 51217 | 14423 | 220010 | 544549 | 10.63 | 2.50 |
| 80 | 0.08489 | 0.34912 | 36794 | 12846 | 151324 | 324540 | 8.82 | 2.46 |
| 85 | 0.13826 | ... | 23948 | 23948 | 173216 | 173216 | 7.23 | 7.23 |

Life Tables: District Vizianagaram

| Age | m(x,n) | q(x,n) | l(x) | d(x,n) | L(x,n) | T(x) | e(x) | a(x,n) |
| --- | --- | --- | --- | --- | --- | --- | --- | --- |
| Male |  |  |  |  |  |  |  |  |
| 0 | 0.04900 | 0.04714 | 100000 | 4714 | 96189 | 6425870 | 64.26 | 0.19 |
| 1 | 0.00083 | 0.00330 | 95286 | 314 | 380343 | 6329681 | 66.43 | 1.45 |
| 5 | 0.00069 | 0.00343 | 94972 | 326 | 474045 | 5949338 | 62.64 | 2.50 |
| 10 | 0.00057 | 0.00286 | 94646 | 271 | 472554 | 5475293 | 57.85 | 2.50 |
| 15 | 0.00135 | 0.00673 | 94375 | 635 | 470453 | 5002739 | 53.01 | 2.76 |
| 20 | 0.00200 | 0.00995 | 93740 | 933 | 466519 | 4532286 | 48.35 | 2.66 |
| 25 | 0.00296 | 0.01470 | 92808 | 1364 | 460781 | 4065768 | 43.81 | 2.61 |
| 30 | 0.00353 | 0.01751 | 91443 | 1602 | 453420 | 3604987 | 39.42 | 2.63 |
| 35 | 0.00571 | 0.02815 | 89842 | 2529 | 443195 | 3151567 | 35.08 | 2.62 |
| 40 | 0.00670 | 0.03296 | 87313 | 2878 | 429593 | 2708372 | 31.02 | 2.58 |
| 45 | 0.00883 | 0.04323 | 84435 | 3651 | 413498 | 2278780 | 26.99 | 2.62 |
| 50 | 0.01319 | 0.06392 | 80785 | 5164 | 391627 | 1865282 | 23.09 | 2.62 |
| 55 | 0.01781 | 0.08542 | 75621 | 6460 | 362732 | 1473655 | 19.49 | 2.62 |
| 60 | 0.02806 | 0.13154 | 69161 | 9097 | 324188 | 1110923 | 16.06 | 2.62 |
| 65 | 0.04269 | 0.19348 | 60064 | 11621 | 272235 | 786735 | 13.10 | 2.58 |
| 70 | 0.06415 | 0.27668 | 48443 | 13403 | 208930 | 514500 | 10.62 | 2.52 |
| 75 | 0.08784 | 0.35845 | 35040 | 12560 | 142987 | 305570 | 8.72 | 2.44 |
| 80 | 0.11322 | 0.43669 | 22480 | 9817 | 86704 | 162583 | 7.23 | 2.38 |
| 85 | 0.16689 | ... | 12663 | 12663 | 75879 | 75879 | 5.99 | 5.99 |
| Female |  |  |  |  |  |  |  |  |
| 0 | 0.04972 | 0.04780 | 100000 | 4780 | 96144 | 6891967 | 68.92 | 0.19 |
| 1 | 0.00128 | 0.00511 | 95220 | 487 | 379637 | 6795823 | 71.37 | 1.45 |
| 5 | 0.00061 | 0.00304 | 94733 | 288 | 472947 | 6416185 | 67.73 | 2.50 |
| 10 | 0.00053 | 0.00266 | 94445 | 251 | 471599 | 5943238 | 62.93 | 2.50 |
| 15 | 0.00158 | 0.00787 | 94194 | 741 | 469305 | 5471639 | 58.09 | 2.75 |
| 20 | 0.00182 | 0.00907 | 93453 | 848 | 465174 | 5002334 | 53.53 | 2.53 |
| 25 | 0.00189 | 0.00939 | 92605 | 870 | 460859 | 4537160 | 48.99 | 2.51 |
| 30 | 0.00192 | 0.00957 | 91736 | 878 | 456499 | 4076301 | 44.44 | 2.52 |
| 35 | 0.00209 | 0.01038 | 90858 | 943 | 452028 | 3619803 | 39.84 | 2.60 |
| 40 | 0.00321 | 0.01595 | 89915 | 1434 | 446165 | 3167774 | 35.23 | 2.62 |
| 45 | 0.00390 | 0.01931 | 88481 | 1709 | 438394 | 2721609 | 30.76 | 2.65 |
| 50 | 0.00698 | 0.03435 | 86772 | 2981 | 427008 | 2283216 | 26.31 | 2.70 |
| 55 | 0.01098 | 0.05355 | 83791 | 4487 | 408624 | 1856207 | 22.15 | 2.70 |
| 60 | 0.02007 | 0.09591 | 79304 | 7606 | 378909 | 1447583 | 18.25 | 2.68 |
| 65 | 0.03253 | 0.15095 | 71698 | 10823 | 332684 | 1068673 | 14.91 | 2.62 |
| 70 | 0.04839 | 0.21648 | 60875 | 13178 | 272357 | 735989 | 12.09 | 2.57 |
| 75 | 0.07395 | 0.31186 | 47697 | 14875 | 201151 | 463632 | 9.72 | 2.49 |
| 80 | 0.09665 | 0.38749 | 32822 | 12718 | 131588 | 262481 | 8.00 | 2.44 |
| 85 | 0.15359 | ... | 20104 | 20104 | 130893 | 130893 | 6.51 | 6.51 |

Life Tables: District Vishakhapatnam

| Age | m(x,n) | q(x,n) | l(x) | d(x,n) | L(x,n) | T(x) | e(x) | a(x,n) |
| --- | --- | --- | --- | --- | --- | --- | --- | --- |
| Male |  |  |  |  |  |  |  |  |
| 0 | 0.04411 | 0.04257 | 100000 | 4257 | 96500 | 6547724 | 65.48 | 0.18 |
| 1 | 0.00047 | 0.00189 | 95743 | 181 | 382512 | 6451224 | 67.38 | 1.45 |
| 5 | 0.00051 | 0.00257 | 95562 | 245 | 477197 | 6068712 | 63.51 | 2.50 |
| 10 | 0.00055 | 0.00276 | 95317 | 263 | 475927 | 5591514 | 58.66 | 2.50 |
| 15 | 0.00118 | 0.00591 | 95054 | 562 | 474006 | 5115588 | 53.82 | 2.75 |
| 20 | 0.00185 | 0.00922 | 94492 | 872 | 470440 | 4641582 | 49.12 | 2.68 |
| 25 | 0.00287 | 0.01426 | 93621 | 1335 | 464926 | 4171142 | 44.55 | 2.62 |
| 30 | 0.00339 | 0.01682 | 92286 | 1552 | 457757 | 3706217 | 40.16 | 2.64 |
| 35 | 0.00569 | 0.02809 | 90733 | 2549 | 447599 | 3248459 | 35.80 | 2.62 |
| 40 | 0.00638 | 0.03140 | 88184 | 2769 | 434208 | 2800860 | 31.76 | 2.58 |
| 45 | 0.00870 | 0.04263 | 85416 | 3641 | 418393 | 2366652 | 27.71 | 2.61 |
| 50 | 0.01205 | 0.05858 | 81775 | 4791 | 397482 | 1948259 | 23.82 | 2.62 |
| 55 | 0.01764 | 0.08467 | 76984 | 6518 | 369494 | 1550777 | 20.14 | 2.63 |
| 60 | 0.02727 | 0.12807 | 70466 | 9024 | 330880 | 1181284 | 16.76 | 2.62 |
| 65 | 0.04190 | 0.19016 | 61441 | 11683 | 278844 | 850403 | 13.84 | 2.57 |
| 70 | 0.05873 | 0.25619 | 49758 | 12747 | 217048 | 571559 | 11.49 | 2.51 |
| 75 | 0.07909 | 0.32899 | 37010 | 12176 | 153955 | 354511 | 9.58 | 2.45 |
| 80 | 0.09999 | 0.39687 | 24834 | 9856 | 98571 | 200556 | 8.08 | 2.40 |
| 85 | 0.14687 | ... | 14978 | 14978 | 101985 | 101985 | 6.81 | 6.81 |
| Female |  |  |  |  |  |  |  |  |
| 0 | 0.04138 | 0.04001 | 100000 | 4001 | 96679 | 7087784 | 70.88 | 0.17 |
| 1 | 0.00074 | 0.00294 | 95999 | 282 | 383279 | 6991105 | 72.82 | 1.46 |
| 5 | 0.00055 | 0.00276 | 95717 | 264 | 477923 | 6607826 | 69.04 | 2.50 |
| 10 | 0.00042 | 0.00209 | 95453 | 199 | 476764 | 6129903 | 64.22 | 2.50 |
| 15 | 0.00115 | 0.00575 | 95253 | 548 | 475039 | 5653138 | 59.35 | 2.76 |
| 20 | 0.00148 | 0.00735 | 94705 | 696 | 471820 | 5178100 | 54.68 | 2.55 |
| 25 | 0.00147 | 0.00732 | 94009 | 688 | 468344 | 4706280 | 50.06 | 2.53 |
| 30 | 0.00169 | 0.00844 | 93321 | 788 | 464704 | 4237936 | 45.41 | 2.59 |
| 35 | 0.00225 | 0.01118 | 92534 | 1035 | 460235 | 3773232 | 40.78 | 2.65 |
| 40 | 0.00353 | 0.01750 | 91499 | 1601 | 453696 | 3312998 | 36.21 | 2.63 |
| 45 | 0.00430 | 0.02128 | 89898 | 1913 | 444939 | 2859301 | 31.81 | 2.62 |
| 50 | 0.00661 | 0.03255 | 87985 | 2864 | 433273 | 2414362 | 27.44 | 2.68 |
| 55 | 0.01079 | 0.05263 | 85121 | 4480 | 415251 | 1981089 | 23.27 | 2.69 |
| 60 | 0.01824 | 0.08746 | 80641 | 7053 | 386678 | 1565838 | 19.42 | 2.66 |
| 65 | 0.02747 | 0.12895 | 73588 | 9489 | 345426 | 1179160 | 16.02 | 2.63 |
| 70 | 0.04423 | 0.19985 | 64099 | 12810 | 289604 | 833734 | 13.01 | 2.59 |
| 75 | 0.06542 | 0.28114 | 51289 | 14419 | 220398 | 544130 | 10.61 | 2.50 |
| 80 | 0.08518 | 0.35012 | 36869 | 12909 | 151547 | 323731 | 8.78 | 2.46 |
| 85 | 0.13916 | ... | 23961 | 23961 | 172185 | 172185 | 7.19 | 7.19 |

Life Tables: District East Godavari

| Age | m(x,n) | q(x,n) | l(x) | d(x,n) | L(x,n) | T(x) | e(x) | a(x,n) |
| --- | --- | --- | --- | --- | --- | --- | --- | --- |
| Male |  |  |  |  |  |  |  |  |
| 0 | 0.03835 | 0.03716 | 100000 | 3716 | 96884 | 6692006 | 66.92 | 0.16 |
| 1 | 0.00028 | 0.00111 | 96284 | 107 | 384866 | 6595122 | 68.50 | 1.46 |
| 5 | 0.00043 | 0.00216 | 96177 | 208 | 480367 | 6210256 | 64.57 | 2.50 |
| 10 | 0.00056 | 0.00281 | 95970 | 270 | 479173 | 5729889 | 59.71 | 2.50 |
| 15 | 0.00106 | 0.00530 | 95700 | 507 | 477338 | 5250716 | 54.87 | 2.71 |
| 20 | 0.00158 | 0.00786 | 95193 | 748 | 474223 | 4773377 | 50.14 | 2.68 |
| 25 | 0.00251 | 0.01246 | 94444 | 1177 | 469454 | 4299154 | 45.52 | 2.65 |
| 30 | 0.00332 | 0.01646 | 93267 | 1535 | 462701 | 3829699 | 41.06 | 2.63 |
| 35 | 0.00490 | 0.02420 | 91732 | 2220 | 453353 | 3366998 | 36.70 | 2.61 |
| 40 | 0.00589 | 0.02903 | 89512 | 2599 | 441287 | 2913645 | 32.55 | 2.59 |
| 45 | 0.00783 | 0.03844 | 86914 | 3341 | 426612 | 2472358 | 28.45 | 2.62 |
| 50 | 0.01127 | 0.05490 | 83572 | 4588 | 406965 | 2045746 | 24.48 | 2.63 |
| 55 | 0.01600 | 0.07705 | 78984 | 6085 | 380439 | 1638781 | 20.75 | 2.62 |
| 60 | 0.02358 | 0.11164 | 72899 | 8138 | 345137 | 1258342 | 17.26 | 2.62 |
| 65 | 0.03631 | 0.16687 | 64760 | 10806 | 297632 | 913205 | 14.10 | 2.58 |
| 70 | 0.04938 | 0.22016 | 53954 | 11878 | 240545 | 615574 | 11.41 | 2.54 |
| 75 | 0.07198 | 0.30544 | 42076 | 12852 | 178547 | 375029 | 8.91 | 2.52 |
| 80 | 0.11335 | 0.43918 | 29224 | 12835 | 113225 | 196482 | 6.72 | 2.44 |
| 85 | 0.19685 | ... | 16390 | 16390 | 83257 | 83257 | 5.08 | 5.08 |
| Female |  |  |  |  |  |  |  |  |
| 0 | 0.02485 | 0.02432 | 100000 | 2432 | 97867 | 7230497 | 72.30 | 0.12 |
| 1 | 0.00091 | 0.00362 | 97568 | 353 | 389384 | 7132630 | 73.10 | 1.48 |
| 5 | 0.00035 | 0.00173 | 97215 | 168 | 485654 | 6743247 | 69.36 | 2.50 |
| 10 | 0.00032 | 0.00158 | 97047 | 153 | 484850 | 6257593 | 64.48 | 2.50 |
| 15 | 0.00066 | 0.00330 | 96893 | 320 | 483729 | 5772743 | 59.58 | 2.69 |
| 20 | 0.00080 | 0.00400 | 96574 | 386 | 481935 | 5289015 | 54.77 | 2.59 |
| 25 | 0.00100 | 0.00500 | 96187 | 481 | 479796 | 4807080 | 49.98 | 2.63 |
| 30 | 0.00150 | 0.00748 | 95706 | 716 | 476809 | 4327284 | 45.21 | 2.59 |
| 35 | 0.00160 | 0.00797 | 94990 | 757 | 473161 | 3850475 | 40.54 | 2.63 |
| 40 | 0.00290 | 0.01439 | 94233 | 1356 | 468077 | 3377314 | 35.84 | 2.72 |
| 45 | 0.00477 | 0.02357 | 92877 | 2189 | 459282 | 2909237 | 31.32 | 2.67 |
| 50 | 0.00681 | 0.03353 | 90688 | 3041 | 446352 | 2449955 | 27.02 | 2.67 |
| 55 | 0.01147 | 0.05588 | 87647 | 4898 | 426890 | 2003603 | 22.86 | 2.68 |
| 60 | 0.01842 | 0.08826 | 82750 | 7303 | 396603 | 1576712 | 19.05 | 2.65 |
| 65 | 0.02867 | 0.13419 | 75446 | 10124 | 353178 | 1180109 | 15.64 | 2.62 |
| 70 | 0.04452 | 0.20109 | 65322 | 13136 | 295063 | 826932 | 12.66 | 2.60 |
| 75 | 0.07173 | 0.30399 | 52186 | 15864 | 221168 | 531868 | 10.19 | 2.49 |
| 80 | 0.08838 | 0.36084 | 36322 | 13107 | 148304 | 310701 | 8.55 | 2.46 |
| 85 | 0.14296 | ... | 23216 | 23216 | 162397 | 162397 | 7.00 | 7.00 |

Life Tables: District West Godavari

| Age | m(x,n) | q(x,n) | l(x) | d(x,n) | L(x,n) | T(x) | e(x) | a(x,n) |
| --- | --- | --- | --- | --- | --- | --- | --- | --- |
| Male |  |  |  |  |  |  |  |  |
| 0 | 0.03796 | 0.03678 | 100000 | 3678 | 96911 | 6704644 | 67.05 | 0.16 |
| 1 | 0.00026 | 0.00104 | 96322 | 100 | 385032 | 6607732 | 68.60 | 1.46 |
| 5 | 0.00043 | 0.00212 | 96221 | 204 | 480595 | 6222700 | 64.67 | 2.50 |
| 10 | 0.00056 | 0.00277 | 96017 | 266 | 479419 | 5742105 | 59.80 | 2.50 |
| 15 | 0.00105 | 0.00523 | 95751 | 501 | 477606 | 5262686 | 54.96 | 2.71 |
| 20 | 0.00154 | 0.00765 | 95250 | 729 | 474554 | 4785080 | 50.24 | 2.67 |
| 25 | 0.00247 | 0.01226 | 94521 | 1159 | 469888 | 4310526 | 45.60 | 2.65 |
| 30 | 0.00331 | 0.01641 | 93362 | 1532 | 463185 | 3840638 | 41.14 | 2.63 |
| 35 | 0.00481 | 0.02375 | 91831 | 2181 | 453936 | 3377453 | 36.78 | 2.61 |
| 40 | 0.00583 | 0.02874 | 89649 | 2576 | 442032 | 2923517 | 32.61 | 2.59 |
| 45 | 0.00775 | 0.03805 | 87073 | 3313 | 427485 | 2481485 | 28.50 | 2.62 |
| 50 | 0.01128 | 0.05493 | 83760 | 4601 | 407880 | 2054000 | 24.52 | 2.63 |
| 55 | 0.01593 | 0.07675 | 79159 | 6075 | 381333 | 1646120 | 20.80 | 2.62 |
| 60 | 0.02345 | 0.11104 | 73084 | 8115 | 346115 | 1264787 | 17.31 | 2.62 |
| 65 | 0.03602 | 0.16562 | 64969 | 10760 | 298775 | 918672 | 14.14 | 2.58 |
| 70 | 0.04869 | 0.21741 | 54209 | 11785 | 242054 | 619897 | 11.44 | 2.54 |
| 75 | 0.07111 | 0.30241 | 42423 | 12829 | 180410 | 377843 | 8.91 | 2.53 |
| 80 | 0.11375 | 0.44048 | 29594 | 13036 | 114596 | 197433 | 6.67 | 2.44 |
| 85 | 0.19989 | ... | 16558 | 16558 | 82837 | 82837 | 5.00 | 5.00 |
| Female |  |  |  |  |  |  |  |  |
| 0 | 0.02451 | 0.02399 | 100000 | 2399 | 97894 | 7241394 | 72.41 | 0.12 |
| 1 | 0.00089 | 0.00355 | 97601 | 346 | 389533 | 7143500 | 73.19 | 1.48 |
| 5 | 0.00034 | 0.00169 | 97255 | 164 | 485862 | 6753968 | 69.45 | 2.50 |
| 10 | 0.00031 | 0.00156 | 97090 | 151 | 485072 | 6268106 | 64.56 | 2.50 |
| 15 | 0.00065 | 0.00324 | 96939 | 314 | 483969 | 5783034 | 59.66 | 2.69 |
| 20 | 0.00079 | 0.00394 | 96625 | 381 | 482204 | 5299065 | 54.84 | 2.59 |
| 25 | 0.00099 | 0.00494 | 96244 | 475 | 480093 | 4816861 | 50.05 | 2.63 |
| 30 | 0.00149 | 0.00742 | 95768 | 711 | 477133 | 4336768 | 45.28 | 2.60 |
| 35 | 0.00159 | 0.00790 | 95058 | 751 | 473513 | 3859635 | 40.60 | 2.63 |
| 40 | 0.00288 | 0.01431 | 94307 | 1350 | 468461 | 3386122 | 35.91 | 2.72 |
| 45 | 0.00474 | 0.02346 | 92957 | 2181 | 459702 | 2917661 | 31.39 | 2.67 |
| 50 | 0.00678 | 0.03335 | 90777 | 3027 | 446826 | 2457959 | 27.08 | 2.67 |
| 55 | 0.01142 | 0.05562 | 87749 | 4881 | 427440 | 2011134 | 22.92 | 2.68 |
| 60 | 0.01832 | 0.08783 | 82869 | 7278 | 397257 | 1583694 | 19.11 | 2.65 |
| 65 | 0.02852 | 0.13353 | 75590 | 10094 | 353972 | 1186437 | 15.70 | 2.62 |
| 70 | 0.04426 | 0.20004 | 65497 | 13102 | 296024 | 832464 | 12.71 | 2.60 |
| 75 | 0.07135 | 0.30265 | 52395 | 15857 | 222235 | 536441 | 10.24 | 2.49 |
| 80 | 0.08778 | 0.35887 | 36537 | 13112 | 149379 | 314205 | 8.60 | 2.46 |
| 85 | 0.14212 | ... | 23425 | 23425 | 164826 | 164826 | 7.04 | 7.04 |

Life Tables: District Krishna

| Age | m(x,n) | q(x,n) | l(x) | d(x,n) | L(x,n) | T(x) | e(x) | a(x,n) |
| --- | --- | --- | --- | --- | --- | --- | --- | --- |
| Male |  |  |  |  |  |  |  |  |
| 0 | 0.04421 | 0.04266 | 100000 | 4266 | 96493 | 6544585 | 65.45 | 0.18 |
| 1 | 0.00048 | 0.00192 | 95734 | 183 | 382468 | 6448092 | 67.35 | 1.45 |
| 5 | 0.00052 | 0.00259 | 95550 | 248 | 477133 | 6065624 | 63.48 | 2.50 |
| 10 | 0.00055 | 0.00277 | 95303 | 264 | 475853 | 5588491 | 58.64 | 2.50 |
| 15 | 0.00119 | 0.00595 | 95039 | 565 | 473921 | 5112638 | 53.80 | 2.75 |
| 20 | 0.00186 | 0.00925 | 94473 | 874 | 470339 | 4638717 | 49.10 | 2.68 |
| 25 | 0.00287 | 0.01428 | 93600 | 1336 | 464818 | 4168378 | 44.53 | 2.62 |
| 30 | 0.00339 | 0.01682 | 92263 | 1552 | 457647 | 3703560 | 40.14 | 2.64 |
| 35 | 0.00570 | 0.02812 | 90711 | 2551 | 447485 | 3245913 | 35.78 | 2.62 |
| 40 | 0.00639 | 0.03144 | 88160 | 2772 | 434080 | 2798429 | 31.74 | 2.57 |
| 45 | 0.00870 | 0.04264 | 85389 | 3641 | 418260 | 2364348 | 27.69 | 2.61 |
| 50 | 0.01208 | 0.05874 | 81748 | 4802 | 397324 | 1946088 | 23.81 | 2.62 |
| 55 | 0.01766 | 0.08475 | 76947 | 6521 | 369296 | 1548764 | 20.13 | 2.63 |
| 60 | 0.02728 | 0.12810 | 70425 | 9021 | 330687 | 1179468 | 16.75 | 2.62 |
| 65 | 0.04196 | 0.19040 | 61404 | 11691 | 278645 | 848780 | 13.82 | 2.57 |
| 70 | 0.05889 | 0.25681 | 49713 | 12767 | 216773 | 570136 | 11.47 | 2.51 |
| 75 | 0.07929 | 0.32969 | 36946 | 12181 | 153614 | 353362 | 9.56 | 2.45 |
| 80 | 0.10018 | 0.39745 | 24765 | 9843 | 98254 | 199748 | 8.07 | 2.40 |
| 85 | 0.14703 | ... | 14922 | 14922 | 101493 | 101493 | 6.80 | 6.80 |
| Female |  |  |  |  |  |  |  |  |
| 0 | 0.04005 | 0.03876 | 100000 | 3876 | 96769 | 7107210 | 71.07 | 0.17 |
| 1 | 0.00069 | 0.00276 | 96124 | 265 | 383822 | 7010441 | 72.93 | 1.46 |
| 5 | 0.00054 | 0.00269 | 95859 | 258 | 478649 | 6626619 | 69.13 | 2.50 |
| 10 | 0.00041 | 0.00206 | 95601 | 197 | 477512 | 6147970 | 64.31 | 2.50 |
| 15 | 0.00106 | 0.00529 | 95404 | 505 | 475880 | 5670458 | 59.44 | 2.74 |
| 20 | 0.00133 | 0.00665 | 94899 | 631 | 472951 | 5194578 | 54.74 | 2.55 |
| 25 | 0.00137 | 0.00685 | 94268 | 646 | 469748 | 4721627 | 50.09 | 2.53 |
| 30 | 0.00159 | 0.00791 | 93622 | 741 | 466331 | 4251879 | 45.42 | 2.59 |
| 35 | 0.00220 | 0.01095 | 92882 | 1017 | 462031 | 3785549 | 40.76 | 2.66 |
| 40 | 0.00352 | 0.01747 | 91865 | 1605 | 455523 | 3323518 | 36.18 | 2.63 |
| 45 | 0.00429 | 0.02125 | 90260 | 1918 | 446742 | 2867994 | 31.77 | 2.62 |
| 50 | 0.00666 | 0.03280 | 88342 | 2898 | 434986 | 2421253 | 27.41 | 2.68 |
| 55 | 0.01087 | 0.05302 | 85444 | 4530 | 416739 | 1986267 | 23.25 | 2.69 |
| 60 | 0.01814 | 0.08701 | 80914 | 7040 | 388026 | 1569528 | 19.40 | 2.65 |
| 65 | 0.02679 | 0.12596 | 73874 | 9305 | 347286 | 1181501 | 15.99 | 2.63 |
| 70 | 0.04360 | 0.19728 | 64569 | 12738 | 292170 | 834216 | 12.92 | 2.59 |
| 75 | 0.06444 | 0.27766 | 51830 | 14391 | 223319 | 542046 | 10.46 | 2.51 |
| 80 | 0.08716 | 0.35688 | 37439 | 13361 | 153295 | 318727 | 8.51 | 2.46 |
| 85 | 0.14555 | ... | 24078 | 24078 | 165432 | 165432 | 6.87 | 6.87 |

Life Tables: District Guntur

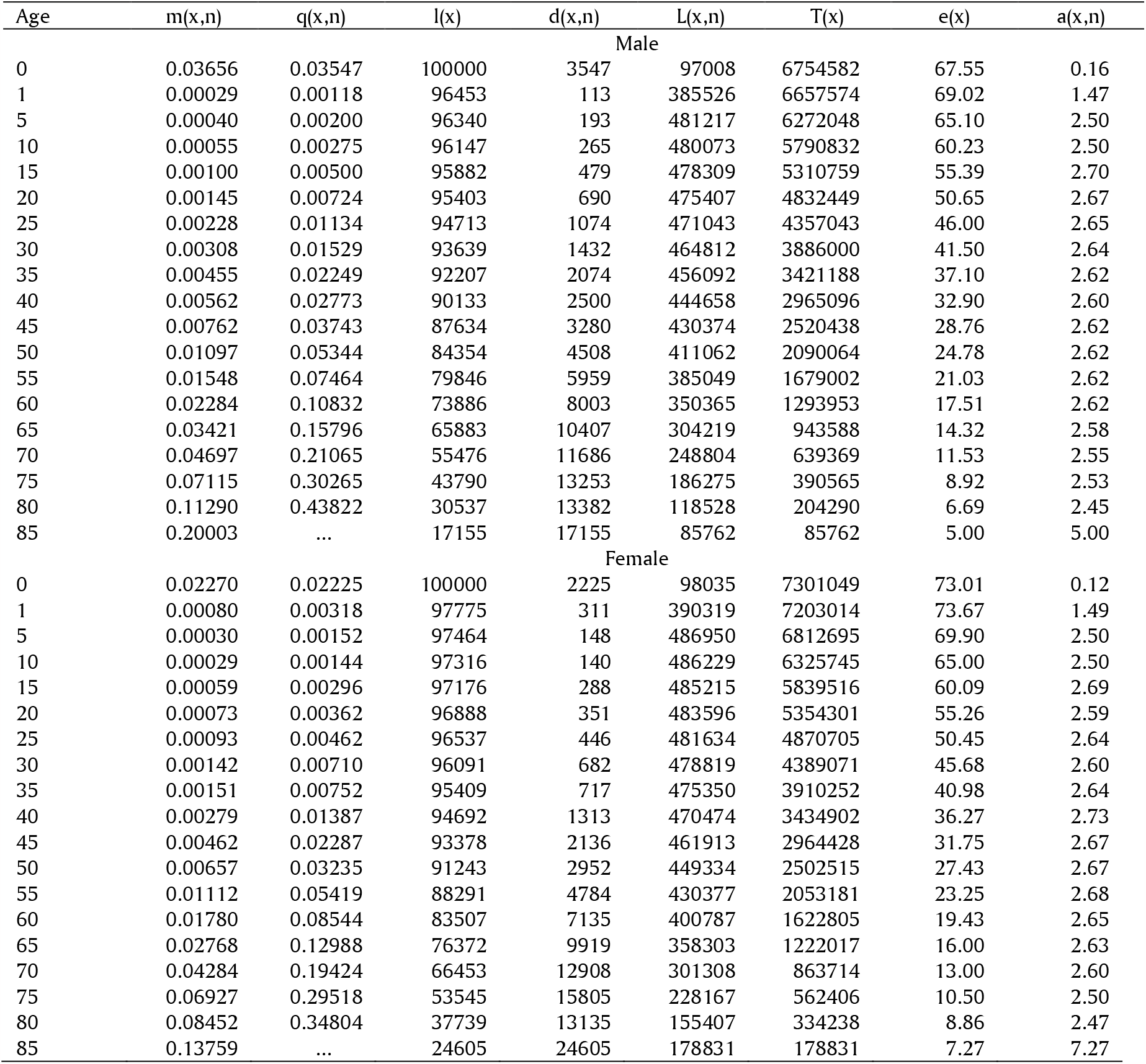

| Age | m(x,n) | q(x,n) | l(x) | d(x,n) | L(x,n) | T(x) | e(x) | a(x,n) |
| --- | --- | --- | --- | --- | --- | --- | --- | --- |
| Male |  |  |  |  |  |  |  |  |
| 0 | 0.03656 | 0.03547 | 100000 | 3547 | 97008 | 6754582 | 67.55 | 0.16 |
| 1 | 0.00029 | 0.00118 | 96453 | 113 | 385526 | 6657574 | 69.02 | 1.47 |
| 5 | 0.00040 | 0.00200 | 96340 | 193 | 481217 | 6272048 | 65.10 | 2.50 |
| 10 | 0.00055 | 0.00275 | 96147 | 265 | 480073 | 5790832 | 60.23 | 2.50 |
| 15 | 0.00100 | 0.00500 | 95882 | 479 | 478309 | 5310759 | 55.39 | 2.70 |
| 20 | 0.00145 | 0.00724 | 95403 | 690 | 475407 | 4832449 | 50.65 | 2.67 |
| 25 | 0.00228 | 0.01134 | 94713 | 1074 | 471043 | 4357043 | 46.00 | 2.65 |
| 30 | 0.00308 | 0.01529 | 93639 | 1432 | 464812 | 3886000 | 41.50 | 2.64 |
| 35 | 0.00455 | 0.02249 | 92207 | 2074 | 456092 | 3421188 | 37.10 | 2.62 |
| 40 | 0.00562 | 0.02773 | 90133 | 2500 | 444658 | 2965096 | 32.90 | 2.60 |
| 45 | 0.00762 | 0.03743 | 87634 | 3280 | 430374 | 2520438 | 28.76 | 2.62 |
| 50 | 0.01097 | 0.05344 | 84354 | 4508 | 411062 | 2090064 | 24.78 | 2.62 |
| 55 | 0.01548 | 0.07464 | 79846 | 5959 | 385049 | 1679002 | 21.03 | 2.62 |
| 60 | 0.02284 | 0.10832 | 73886 | 8003 | 350365 | 1293953 | 17.51 | 2.62 |
| 65 | 0.03421 | 0.15796 | 65883 | 10407 | 304219 | 943588 | 14.32 | 2.58 |
| 70 | 0.04697 | 0.21065 | 55476 | 11686 | 248804 | 639369 | 11.53 | 2.55 |
| 75 | 0.07115 | 0.30265 | 43790 | 13253 | 186275 | 390565 | 8.92 | 2.53 |
| 80 | 0.11290 | 0.43822 | 30537 | 13382 | 118528 | 204290 | 6.69 | 2.45 |
| 85 | 0.20003 | ... | 17155 | 17155 | 85762 | 85762 | 5.00 | 5.00 |
| Female |  |  |  |  |  |  |  |  |
| 0 | 0.02270 | 0.02225 | 100000 | 2225 | 98035 | 7301049 | 73.01 | 0.12 |
| 1 | 0.00080 | 0.00318 | 97775 | 311 | 390319 | 7203014 | 73.67 | 1.49 |
| 5 | 0.00030 | 0.00152 | 97464 | 148 | 486950 | 6812695 | 69.90 | 2.50 |
| 10 | 0.00029 | 0.00144 | 97316 | 140 | 486229 | 6325745 | 65.00 | 2.50 |
| 15 | 0.00059 | 0.00296 | 97176 | 288 | 485215 | 5839516 | 60.09 | 2.69 |
| 20 | 0.00073 | 0.00362 | 96888 | 351 | 483596 | 5354301 | 55.26 | 2.59 |
| 25 | 0.00093 | 0.00462 | 96537 | 446 | 481634 | 4870705 | 50.45 | 2.64 |
| 30 | 0.00142 | 0.00710 | 96091 | 682 | 478819 | 4389071 | 45.68 | 2.60 |
| 35 | 0.00151 | 0.00752 | 95409 | 717 | 475350 | 3910252 | 40.98 | 2.64 |
| 40 | 0.00279 | 0.01387 | 94692 | 1313 | 470474 | 3434902 | 36.27 | 2.73 |
| 45 | 0.00462 | 0.02287 | 93378 | 2136 | 461913 | 2964428 | 31.75 | 2.67 |
| 50 | 0.00657 | 0.03235 | 91243 | 2952 | 449334 | 2502515 | 27.43 | 2.67 |
| 55 | 0.01112 | 0.05419 | 88291 | 4784 | 430377 | 2053181 | 23.25 | 2.68 |
| 60 | 0.01780 | 0.08544 | 83507 | 7135 | 400787 | 1622805 | 19.43 | 2.65 |
| 65 | 0.02768 | 0.12988 | 76372 | 9919 | 358303 | 1222017 | 16.00 | 2.63 |
| 70 | 0.04284 | 0.19424 | 66453 | 12908 | 301308 | 863714 | 13.00 | 2.60 |
| 75 | 0.06927 | 0.29518 | 53545 | 15805 | 228167 | 562406 | 10.50 | 2.50 |
| 80 | 0.08452 | 0.34804 | 37739 | 13135 | 155407 | 334238 | 8.86 | 2.47 |
| 85 | 0.13759 | ... | 24605 | 24605 | 178831 | 178831 | 7.27 | 7.27 |

Life Tables: District Prakasam

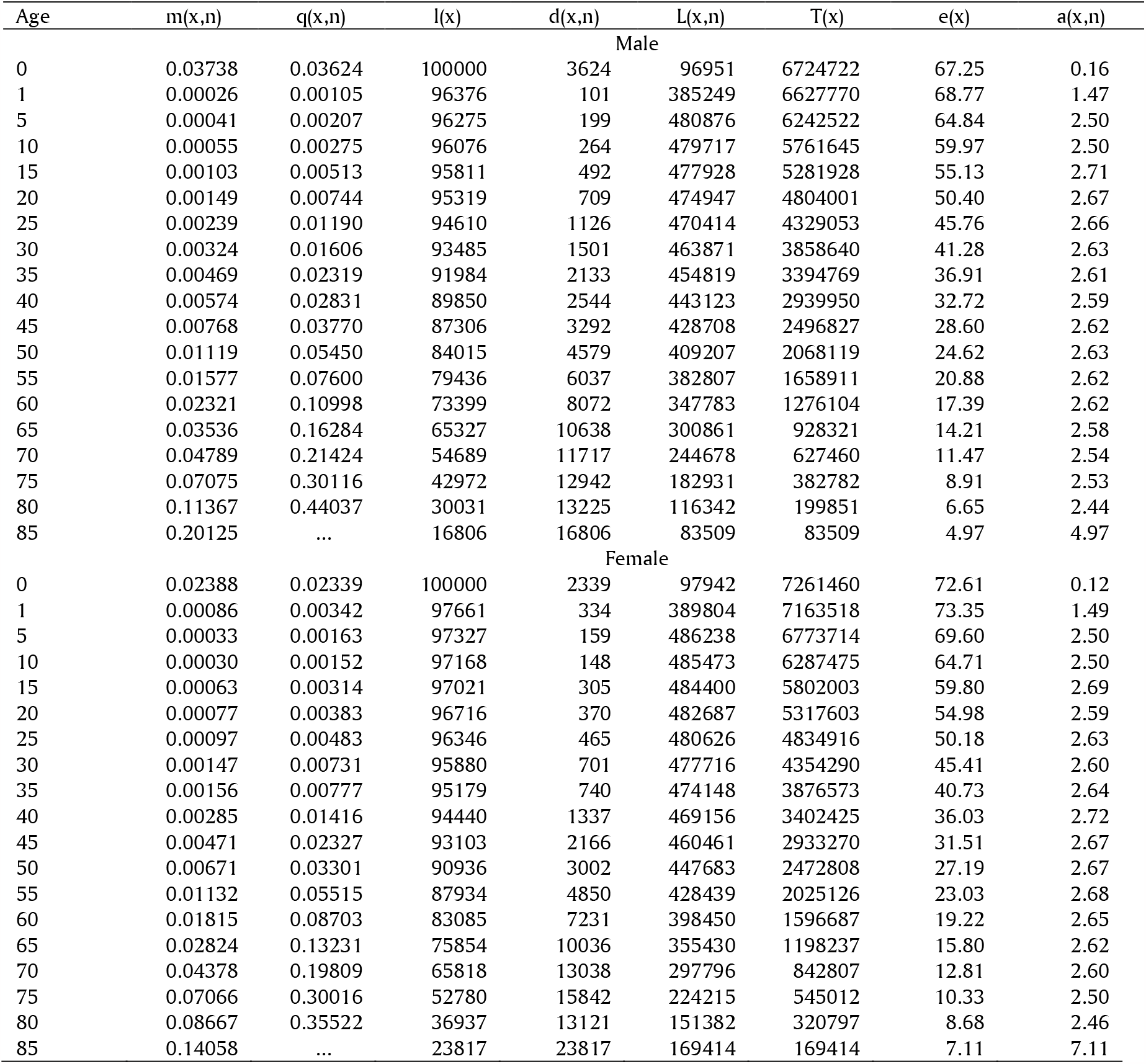

| Age | m(x,n) | q(x,n) | l(x) | d(x,n) | L(x,n) | T(x) | e(x) | a(x,n) |
| --- | --- | --- | --- | --- | --- | --- | --- | --- |
| Male |  |  |  |  |  |  |  |  |
| 0 | 0.03738 | 0.03624 | 100000 | 3624 | 96951 | 6724722 | 67.25 | 0.16 |
| 1 | 0.00026 | 0.00105 | 96376 | 101 | 385249 | 6627770 | 68.77 | 1.47 |
| 5 | 0.00041 | 0.00207 | 96275 | 199 | 480876 | 6242522 | 64.84 | 2.50 |
| 10 | 0.00055 | 0.00275 | 96076 | 264 | 479717 | 5761645 | 59.97 | 2.50 |
| 15 | 0.00103 | 0.00513 | 95811 | 492 | 477928 | 5281928 | 55.13 | 2.71 |
| 20 | 0.00149 | 0.00744 | 95319 | 709 | 474947 | 4804001 | 50.40 | 2.67 |
| 25 | 0.00239 | 0.01190 | 94610 | 1126 | 470414 | 4329053 | 45.76 | 2.66 |
| 30 | 0.00324 | 0.01606 | 93485 | 1501 | 463871 | 3858640 | 41.28 | 2.63 |
| 35 | 0.00469 | 0.02319 | 91984 | 2133 | 454819 | 3394769 | 36.91 | 2.61 |
| 40 | 0.00574 | 0.02831 | 89850 | 2544 | 443123 | 2939950 | 32.72 | 2.59 |
| 45 | 0.00768 | 0.03770 | 87306 | 3292 | 428708 | 2496827 | 28.60 | 2.62 |
| 50 | 0.01119 | 0.05450 | 84015 | 4579 | 409207 | 2068119 | 24.62 | 2.63 |
| 55 | 0.01577 | 0.07600 | 79436 | 6037 | 382807 | 1658911 | 20.88 | 2.62 |
| 60 | 0.02321 | 0.10998 | 73399 | 8072 | 347783 | 1276104 | 17.39 | 2.62 |
| 65 | 0.03536 | 0.16284 | 65327 | 10638 | 300861 | 928321 | 14.21 | 2.58 |
| 70 | 0.04789 | 0.21424 | 54689 | 11717 | 244678 | 627460 | 11.47 | 2.54 |
| 75 | 0.07075 | 0.30116 | 42972 | 12942 | 182931 | 382782 | 8.91 | 2.53 |
| 80 | 0.11367 | 0.44037 | 30031 | 13225 | 116342 | 199851 | 6.65 | 2.44 |
| 85 | 0.20125 | ... | 16806 | 16806 | 83509 | 83509 | 4.97 | 4.97 |
| Female |  |  |  |  |  |  |  |  |
| 0 | 0.02388 | 0.02339 | 100000 | 2339 | 97942 | 7261460 | 72.61 | 0.12 |
| 1 | 0.00086 | 0.00342 | 97661 | 334 | 389804 | 7163518 | 73.35 | 1.49 |
| 5 | 0.00033 | 0.00163 | 97327 | 159 | 486238 | 6773714 | 69.60 | 2.50 |
| 10 | 0.00030 | 0.00152 | 97168 | 148 | 485473 | 6287475 | 64.71 | 2.50 |
| 15 | 0.00063 | 0.00314 | 97021 | 305 | 484400 | 5802003 | 59.80 | 2.69 |
| 20 | 0.00077 | 0.00383 | 96716 | 370 | 482687 | 5317603 | 54.98 | 2.59 |
| 25 | 0.00097 | 0.00483 | 96346 | 465 | 480626 | 4834916 | 50.18 | 2.63 |
| 30 | 0.00147 | 0.00731 | 95880 | 701 | 477716 | 4354290 | 45.41 | 2.60 |
| 35 | 0.00156 | 0.00777 | 95179 | 740 | 474148 | 3876573 | 40.73 | 2.64 |
| 40 | 0.00285 | 0.01416 | 94440 | 1337 | 469156 | 3402425 | 36.03 | 2.72 |
| 45 | 0.00471 | 0.02327 | 93103 | 2166 | 460461 | 2933270 | 31.51 | 2.67 |
| 50 | 0.00671 | 0.03301 | 90936 | 3002 | 447683 | 2472808 | 27.19 | 2.67 |
| 55 | 0.01132 | 0.05515 | 87934 | 4850 | 428439 | 2025126 | 23.03 | 2.68 |
| 60 | 0.01815 | 0.08703 | 83085 | 7231 | 398450 | 1596687 | 19.22 | 2.65 |
| 65 | 0.02824 | 0.13231 | 75854 | 10036 | 355430 | 1198237 | 15.80 | 2.62 |
| 70 | 0.04378 | 0.19809 | 65818 | 13038 | 297796 | 842807 | 12.81 | 2.60 |
| 75 | 0.07066 | 0.30016 | 52780 | 15842 | 224215 | 545012 | 10.33 | 2.50 |
| 80 | 0.08667 | 0.35522 | 36937 | 13121 | 151382 | 320797 | 8.68 | 2.46 |
| 85 | 0.14058 | ... | 23817 | 23817 | 169414 | 169414 | 7.11 | 7.11 |

Life Tables: District Sri Potti Sriramulu Nellore

| Age | m(x,n) | q(x,n) | l(x) | d(x,n) | L(x,n) | T(x) | e(x) | a(x,n) |
| --- | --- | --- | --- | --- | --- | --- | --- | --- |
| Male |  |  |  |  |  |  |  |  |
| 0 | 0.03604 | 0.03497 | 100000 | 3497 | 97044 | 6773884 | 67.74 | 0.15 |
| 1 | 0.00031 | 0.00125 | 96503 | 121 | 385703 | 6676840 | 69.19 | 1.47 |
| 5 | 0.00039 | 0.00196 | 96381 | 189 | 481435 | 6291136 | 65.27 | 2.50 |
| 10 | 0.00055 | 0.00275 | 96193 | 265 | 480301 | 5809701 | 60.40 | 2.50 |
| 15 | 0.00098 | 0.00491 | 95928 | 471 | 478555 | 5329400 | 55.56 | 2.70 |
| 20 | 0.00143 | 0.00711 | 95457 | 678 | 475701 | 4850845 | 50.82 | 2.67 |
| 25 | 0.00221 | 0.01098 | 94779 | 1041 | 471446 | 4375144 | 46.16 | 2.65 |
| 30 | 0.00298 | 0.01481 | 93738 | 1388 | 465412 | 3903698 | 41.64 | 2.64 |
| 35 | 0.00445 | 0.02204 | 92350 | 2035 | 456903 | 3438286 | 37.23 | 2.62 |
| 40 | 0.00554 | 0.02736 | 90314 | 2471 | 445638 | 2981382 | 33.01 | 2.60 |
| 45 | 0.00758 | 0.03725 | 87843 | 3272 | 431440 | 2535744 | 28.87 | 2.62 |
| 50 | 0.01083 | 0.05277 | 84571 | 4463 | 412251 | 2104304 | 24.88 | 2.62 |
| 55 | 0.01529 | 0.07376 | 80109 | 5909 | 386489 | 1692053 | 21.12 | 2.62 |
| 60 | 0.02261 | 0.10725 | 74200 | 7958 | 352028 | 1305564 | 17.60 | 2.62 |
| 65 | 0.03348 | 0.15485 | 66242 | 10258 | 306384 | 953536 | 14.39 | 2.58 |
| 70 | 0.04638 | 0.20835 | 55984 | 11664 | 251472 | 647152 | 11.56 | 2.56 |
| 75 | 0.07138 | 0.30351 | 44320 | 13452 | 188450 | 395680 | 8.93 | 2.54 |
| 80 | 0.11239 | 0.43678 | 30868 | 13483 | 119964 | 207230 | 6.71 | 2.45 |
| 85 | 0.19923 | ... | 17386 | 17386 | 87266 | 87266 | 5.02 | 5.02 |
| Female |  |  |  |  |  |  |  |  |
| 0 | 0.02181 | 0.02140 | 100000 | 2140 | 98104 | 7332246 | 73.32 | 0.11 |
| 1 | 0.00075 | 0.00301 | 97860 | 295 | 390700 | 7234142 | 73.92 | 1.49 |
| 5 | 0.00029 | 0.00144 | 97565 | 140 | 487476 | 6843441 | 70.14 | 2.50 |
| 10 | 0.00028 | 0.00138 | 97425 | 134 | 486789 | 6355965 | 65.24 | 2.50 |
| 15 | 0.00056 | 0.00282 | 97291 | 274 | 485819 | 5869177 | 60.33 | 2.69 |
| 20 | 0.00070 | 0.00347 | 97016 | 337 | 484271 | 5383358 | 55.49 | 2.59 |
| 25 | 0.00089 | 0.00446 | 96679 | 431 | 482381 | 4899087 | 50.67 | 2.64 |
| 30 | 0.00139 | 0.00694 | 96248 | 668 | 479639 | 4416706 | 45.89 | 2.60 |
| 35 | 0.00147 | 0.00732 | 95580 | 700 | 476249 | 3937067 | 41.19 | 2.64 |
| 40 | 0.00274 | 0.01364 | 94881 | 1294 | 471466 | 3460818 | 36.48 | 2.73 |
| 45 | 0.00456 | 0.02256 | 93587 | 2111 | 463011 | 2989352 | 31.94 | 2.67 |
| 50 | 0.00646 | 0.03182 | 91475 | 2911 | 450592 | 2526341 | 27.62 | 2.67 |
| 55 | 0.01096 | 0.05343 | 88564 | 4732 | 431868 | 2075749 | 23.44 | 2.69 |
| 60 | 0.01753 | 0.08417 | 83832 | 7056 | 402603 | 1643881 | 19.61 | 2.65 |
| 65 | 0.02725 | 0.12795 | 76776 | 9824 | 360558 | 1241278 | 16.17 | 2.63 |
| 70 | 0.04210 | 0.19119 | 66953 | 12801 | 304085 | 880719 | 13.15 | 2.60 |
| 75 | 0.06816 | 0.29116 | 54152 | 15767 | 231327 | 576634 | 10.65 | 2.50 |
| 80 | 0.08283 | 0.34236 | 38385 | 13142 | 158657 | 345307 | 9.00 | 2.47 |
| 85 | 0.13525 | ... | 25244 | 25244 | 186650 | 186650 | 7.39 | 7.39 |

Life Tables: District Y.S.R.

| Age | m(x,n) | q(x,n) | l(x) | d(x,n) | L(x,n) | T(x) | e(x) | a(x,n) |
| --- | --- | --- | --- | --- | --- | --- | --- | --- |
| Male |  |  |  |  |  |  |  |  |
| 0 | 0.03781 | 0.03665 | 100000 | 3665 | 96921 | 6709302 | 67.09 | 0.16 |
| 1 | 0.00025 | 0.00102 | 96335 | 98 | 385093 | 6612381 | 68.64 | 1.46 |
| 5 | 0.00042 | 0.00211 | 96237 | 203 | 480679 | 6227288 | 64.71 | 2.50 |
| 10 | 0.00055 | 0.00276 | 96034 | 265 | 479508 | 5746609 | 59.84 | 2.50 |
| 15 | 0.00104 | 0.00521 | 95769 | 499 | 477703 | 5267101 | 55.00 | 2.71 |
| 20 | 0.00152 | 0.00757 | 95270 | 722 | 474674 | 4789398 | 50.27 | 2.67 |
| 25 | 0.00245 | 0.01218 | 94549 | 1152 | 470045 | 4314724 | 45.63 | 2.66 |
| 30 | 0.00330 | 0.01639 | 93397 | 1530 | 463361 | 3844679 | 41.16 | 2.63 |
| 35 | 0.00477 | 0.02359 | 91867 | 2167 | 454148 | 3381317 | 36.81 | 2.61 |
| 40 | 0.00581 | 0.02863 | 89699 | 2568 | 442303 | 2927170 | 32.63 | 2.59 |
| 45 | 0.00772 | 0.03791 | 87131 | 3303 | 427802 | 2484867 | 28.52 | 2.62 |
| 50 | 0.01128 | 0.05493 | 83828 | 4605 | 408214 | 2057065 | 24.54 | 2.63 |
| 55 | 0.01591 | 0.07663 | 79223 | 6071 | 381661 | 1648851 | 20.81 | 2.62 |
| 60 | 0.02340 | 0.11081 | 73152 | 8106 | 346476 | 1267190 | 17.32 | 2.62 |
| 65 | 0.03591 | 0.16517 | 65046 | 10744 | 299197 | 920714 | 14.15 | 2.58 |
| 70 | 0.04844 | 0.21641 | 54303 | 11752 | 242610 | 621516 | 11.45 | 2.54 |
| 75 | 0.07080 | 0.30130 | 42551 | 12821 | 181095 | 378906 | 8.90 | 2.53 |
| 80 | 0.11388 | 0.44091 | 29730 | 13108 | 115105 | 197811 | 6.65 | 2.44 |
| 85 | 0.20098 | ... | 16622 | 16622 | 82706 | 82706 | 4.98 | 4.98 |
| Female |  |  |  |  |  |  |  |  |
| 0 | 0.02370 | 0.02322 | 100000 | 2322 | 97956 | 7267052 | 72.67 | 0.12 |
| 1 | 0.00085 | 0.00339 | 97678 | 331 | 389880 | 7169096 | 73.40 | 1.49 |
| 5 | 0.00032 | 0.00162 | 97347 | 158 | 486340 | 6779217 | 69.64 | 2.50 |
| 10 | 0.00030 | 0.00151 | 97189 | 147 | 485579 | 6292877 | 64.75 | 2.50 |
| 15 | 0.00062 | 0.00312 | 97042 | 303 | 484513 | 5807298 | 59.84 | 2.69 |
| 20 | 0.00076 | 0.00380 | 96740 | 368 | 482812 | 5322785 | 55.02 | 2.59 |
| 25 | 0.00096 | 0.00480 | 96372 | 463 | 480766 | 4839973 | 50.22 | 2.63 |
| 30 | 0.00146 | 0.00728 | 95909 | 698 | 477869 | 4359207 | 45.45 | 2.60 |
| 35 | 0.00155 | 0.00773 | 95211 | 736 | 474316 | 3881338 | 40.77 | 2.64 |
| 40 | 0.00284 | 0.01412 | 94475 | 1334 | 469341 | 3407023 | 36.06 | 2.72 |
| 45 | 0.00469 | 0.02321 | 93141 | 2162 | 460666 | 2937682 | 31.54 | 2.67 |
| 50 | 0.00669 | 0.03292 | 90979 | 2995 | 447916 | 2477016 | 27.23 | 2.67 |
| 55 | 0.01129 | 0.05501 | 87984 | 4840 | 428712 | 2029100 | 23.06 | 2.68 |
| 60 | 0.01810 | 0.08680 | 83144 | 7217 | 398782 | 1600388 | 19.25 | 2.65 |
| 65 | 0.02816 | 0.13196 | 75927 | 10019 | 355839 | 1201607 | 15.83 | 2.62 |
| 70 | 0.04365 | 0.19754 | 65908 | 13019 | 298296 | 845768 | 12.83 | 2.60 |
| 75 | 0.07046 | 0.29945 | 52889 | 15837 | 224776 | 547472 | 10.35 | 2.50 |
| 80 | 0.08636 | 0.35418 | 37051 | 13123 | 151953 | 322696 | 8.71 | 2.46 |
| 85 | 0.14014 | ... | 23928 | 23928 | 170743 | 170743 | 7.14 | 7.14 |

Life Tables: District Kurnool

| Age | m(x,n) | q(x,n) | l(x) | d(x,n) | L(x,n) | T(x) | e(x) | a(x,n) |
| --- | --- | --- | --- | --- | --- | --- | --- | --- |
| Male |  |  |  |  |  |  |  |  |
| 0 | 0.04002 | 0.03873 | 100000 | 3873 | 96771 | 6640740 | 66.41 | 0.17 |
| 1 | 0.00036 | 0.00143 | 96127 | 137 | 384161 | 6543970 | 68.08 | 1.46 |
| 5 | 0.00047 | 0.00232 | 95990 | 223 | 479393 | 6159809 | 64.17 | 2.50 |
| 10 | 0.00060 | 0.00297 | 95767 | 285 | 478124 | 5680416 | 59.31 | 2.50 |
| 15 | 0.00112 | 0.00559 | 95482 | 534 | 476196 | 5202293 | 54.48 | 2.72 |
| 20 | 0.00177 | 0.00880 | 94948 | 836 | 472802 | 4726096 | 49.78 | 2.68 |
| 25 | 0.00269 | 0.01336 | 94113 | 1257 | 467581 | 4253294 | 45.19 | 2.63 |
| 30 | 0.00336 | 0.01665 | 92855 | 1546 | 460619 | 3785714 | 40.77 | 2.63 |
| 35 | 0.00529 | 0.02614 | 91309 | 2387 | 450856 | 3325095 | 36.42 | 2.62 |
| 40 | 0.00615 | 0.03029 | 88923 | 2693 | 438090 | 2874239 | 32.32 | 2.58 |
| 45 | 0.00817 | 0.04009 | 86230 | 3457 | 422880 | 2436149 | 28.25 | 2.61 |
| 50 | 0.01121 | 0.05459 | 82773 | 4518 | 403109 | 2013269 | 24.32 | 2.62 |
| 55 | 0.01623 | 0.07813 | 78255 | 6114 | 376756 | 1610161 | 20.58 | 2.63 |
| 60 | 0.02410 | 0.11399 | 72140 | 8223 | 341166 | 1233404 | 17.10 | 2.62 |
| 65 | 0.03750 | 0.17193 | 63917 | 10990 | 293031 | 892238 | 13.96 | 2.58 |
| 70 | 0.05237 | 0.23193 | 52928 | 12276 | 234406 | 599207 | 11.32 | 2.54 |
| 75 | 0.07569 | 0.31820 | 40652 | 12935 | 170906 | 364801 | 8.97 | 2.50 |
| 80 | 0.11096 | 0.43151 | 27717 | 11960 | 107789 | 193895 | 7.00 | 2.43 |
| 85 | 0.18299 | ... | 15757 | 15757 | 86106 | 86106 | 5.46 | 5.46 |
| Female |  |  |  |  |  |  |  |  |
| 0 | 0.03892 | 0.03769 | 100000 | 3769 | 96846 | 7119840 | 71.20 | 0.16 |
| 1 | 0.00068 | 0.00273 | 96231 | 263 | 384257 | 7022995 | 72.98 | 1.46 |
| 5 | 0.00053 | 0.00263 | 95968 | 252 | 479210 | 6638737 | 69.18 | 2.50 |
| 10 | 0.00039 | 0.00197 | 95716 | 189 | 478108 | 6159527 | 64.35 | 2.50 |
| 15 | 0.00100 | 0.00501 | 95527 | 479 | 476554 | 5681419 | 59.47 | 2.74 |
| 20 | 0.00125 | 0.00622 | 95049 | 591 | 473797 | 5204865 | 54.76 | 2.55 |
| 25 | 0.00131 | 0.00655 | 94458 | 619 | 470766 | 4731067 | 50.09 | 2.54 |
| 30 | 0.00153 | 0.00764 | 93839 | 717 | 467476 | 4260301 | 45.40 | 2.60 |
| 35 | 0.00219 | 0.01089 | 93122 | 1014 | 463247 | 3792826 | 40.73 | 2.67 |
| 40 | 0.00355 | 0.01761 | 92108 | 1622 | 456700 | 3329579 | 36.15 | 2.63 |
| 45 | 0.00430 | 0.02127 | 90486 | 1925 | 447854 | 2872879 | 31.75 | 2.62 |
| 50 | 0.00669 | 0.03294 | 88561 | 2917 | 436038 | 2425024 | 27.38 | 2.68 |
| 55 | 0.01090 | 0.05314 | 85644 | 4551 | 417680 | 1988987 | 23.22 | 2.68 |
| 60 | 0.01805 | 0.08658 | 81093 | 7021 | 388951 | 1571307 | 19.38 | 2.65 |
| 65 | 0.02657 | 0.12495 | 74072 | 9255 | 348397 | 1182356 | 15.96 | 2.63 |
| 70 | 0.04332 | 0.19615 | 64817 | 12714 | 293480 | 833959 | 12.87 | 2.59 |
| 75 | 0.06400 | 0.27608 | 52103 | 14385 | 224770 | 540480 | 10.37 | 2.52 |
| 80 | 0.08835 | 0.36089 | 37718 | 13612 | 154076 | 315709 | 8.37 | 2.46 |
| 85 | 0.14914 | ... | 24106 | 24106 | 161633 | 161633 | 6.71 | 6.71 |

Life Tables: District Anantapur

| Age | m(x,n) | q(x,n) | l(x) | d(x,n) | L(x,n) | T(x) | e(x) | a(x,n) |
| --- | --- | --- | --- | --- | --- | --- | --- | --- |
| Male |  |  |  |  |  |  |  |  |
| 0 | 0.04630 | 0.04461 | 100000 | 4461 | 96359 | 6483606 | 64.84 | 0.18 |
| 1 | 0.00059 | 0.00236 | 95539 | 226 | 381579 | 6387247 | 66.86 | 1.45 |
| 5 | 0.00063 | 0.00315 | 95313 | 300 | 475815 | 6005669 | 63.01 | 2.50 |
| 10 | 0.00060 | 0.00301 | 95013 | 286 | 474350 | 5529854 | 58.20 | 2.50 |
| 15 | 0.00137 | 0.00683 | 94727 | 647 | 472174 | 5055504 | 53.37 | 2.74 |
| 20 | 0.00195 | 0.00969 | 94080 | 911 | 468262 | 4583330 | 48.72 | 2.65 |
| 25 | 0.00293 | 0.01456 | 93169 | 1356 | 462601 | 4115068 | 44.17 | 2.61 |
| 30 | 0.00339 | 0.01683 | 91812 | 1545 | 455407 | 3652467 | 39.78 | 2.64 |
| 35 | 0.00581 | 0.02867 | 90267 | 2588 | 445188 | 3197060 | 35.42 | 2.62 |
| 40 | 0.00655 | 0.03221 | 87679 | 2824 | 431534 | 2751871 | 31.39 | 2.57 |
| 45 | 0.00871 | 0.04269 | 84855 | 3622 | 415655 | 2320337 | 27.34 | 2.62 |
| 50 | 0.01274 | 0.06184 | 81233 | 5024 | 394228 | 1904682 | 23.45 | 2.62 |
| 55 | 0.01797 | 0.08615 | 76209 | 6566 | 365427 | 1510455 | 19.82 | 2.62 |
| 60 | 0.02731 | 0.12824 | 69643 | 8931 | 327007 | 1145027 | 16.44 | 2.63 |
| 65 | 0.04307 | 0.19505 | 60712 | 11842 | 274926 | 818020 | 13.47 | 2.58 |
| 70 | 0.06223 | 0.26939 | 48870 | 13165 | 211546 | 543094 | 11.11 | 2.51 |
| 75 | 0.08345 | 0.34365 | 35705 | 12270 | 147028 | 331548 | 9.29 | 2.43 |
| 80 | 0.10387 | 0.40860 | 23435 | 9576 | 92187 | 184520 | 7.87 | 2.39 |
| 85 | 0.15010 | ... | 13860 | 13860 | 92333 | 92333 | 6.66 | 6.66 |
| Female |  |  |  |  |  |  |  |  |
| 0 | 0.04946 | 0.04756 | 100000 | 4756 | 96160 | 6898991 | 68.99 | 0.19 |
| 1 | 0.00126 | 0.00501 | 95244 | 477 | 379758 | 6802831 | 71.43 | 1.45 |
| 5 | 0.00061 | 0.00305 | 94767 | 289 | 473112 | 6423073 | 67.78 | 2.50 |
| 10 | 0.00053 | 0.00263 | 94478 | 248 | 471768 | 5949961 | 62.98 | 2.50 |
| 15 | 0.00156 | 0.00778 | 94229 | 733 | 469500 | 5478194 | 58.14 | 2.75 |
| 20 | 0.00182 | 0.00904 | 93496 | 845 | 465398 | 5008693 | 53.57 | 2.54 |
| 25 | 0.00189 | 0.00939 | 92651 | 870 | 461085 | 4543295 | 49.04 | 2.51 |
| 30 | 0.00191 | 0.00949 | 91781 | 871 | 456743 | 4082210 | 44.48 | 2.52 |
| 35 | 0.00210 | 0.01043 | 90910 | 948 | 452280 | 3625466 | 39.88 | 2.61 |
| 40 | 0.00323 | 0.01603 | 89962 | 1442 | 446382 | 3173187 | 35.27 | 2.62 |
| 45 | 0.00391 | 0.01938 | 88520 | 1716 | 438568 | 2726805 | 30.80 | 2.65 |
| 50 | 0.00691 | 0.03403 | 86804 | 2954 | 427227 | 2288237 | 26.36 | 2.70 |
| 55 | 0.01096 | 0.05343 | 83850 | 4480 | 408942 | 1861010 | 22.19 | 2.70 |
| 60 | 0.02004 | 0.09576 | 79370 | 7600 | 379254 | 1452068 | 18.29 | 2.68 |
| 65 | 0.03249 | 0.15078 | 71770 | 10821 | 333048 | 1072814 | 14.95 | 2.62 |
| 70 | 0.04835 | 0.21634 | 60948 | 13186 | 272702 | 739766 | 12.14 | 2.57 |
| 75 | 0.07377 | 0.31121 | 47763 | 14864 | 201484 | 467065 | 9.78 | 2.49 |
| 80 | 0.09576 | 0.38461 | 32898 | 12653 | 132137 | 265580 | 8.07 | 2.44 |
| 85 | 0.15171 | ... | 20245 | 20245 | 133444 | 133444 | 6.59 | 6.59 |

Life Tables: District Chittoor

| Age | m(x,n) | q(x,n) | l(x) | d(x,n) | L(x,n) | T(x) | e(x) | a(x,n) |
| --- | --- | --- | --- | --- | --- | --- | --- | --- |
| Male |  |  |  |  |  |  |  |  |
| 0 | 0.04176 | 0.04036 | 100000 | 4036 | 96654 | 6598835 | 65.99 | 0.17 |
| 1 | 0.00041 | 0.00163 | 95964 | 157 | 383457 | 6502180 | 67.76 | 1.46 |
| 5 | 0.00049 | 0.00243 | 95807 | 233 | 478453 | 6118723 | 63.87 | 2.50 |
| 10 | 0.00058 | 0.00291 | 95574 | 278 | 477177 | 5640270 | 59.01 | 2.50 |
| 15 | 0.00115 | 0.00574 | 95297 | 547 | 475245 | 5163093 | 54.18 | 2.74 |
| 20 | 0.00182 | 0.00906 | 94750 | 859 | 471758 | 4687848 | 49.48 | 2.68 |
| 25 | 0.00278 | 0.01380 | 93891 | 1296 | 466375 | 4216090 | 44.90 | 2.62 |
| 30 | 0.00337 | 0.01674 | 92595 | 1550 | 459310 | 3749715 | 40.50 | 2.63 |
| 35 | 0.00549 | 0.02710 | 91045 | 2467 | 449348 | 3290405 | 36.14 | 2.62 |
| 40 | 0.00626 | 0.03085 | 88578 | 2733 | 436266 | 2841058 | 32.07 | 2.58 |
| 45 | 0.00842 | 0.04125 | 85845 | 3541 | 420761 | 2404791 | 28.01 | 2.61 |
| 50 | 0.01152 | 0.05607 | 82304 | 4615 | 400538 | 1984030 | 24.11 | 2.62 |
| 55 | 0.01680 | 0.08077 | 77689 | 6275 | 373572 | 1583493 | 20.38 | 2.63 |
| 60 | 0.02538 | 0.11969 | 71414 | 8547 | 336765 | 1209921 | 16.94 | 2.62 |
| 65 | 0.03931 | 0.17948 | 62867 | 11284 | 287024 | 873156 | 13.89 | 2.58 |
| 70 | 0.05510 | 0.24244 | 51583 | 12506 | 226980 | 586132 | 11.36 | 2.53 |
| 75 | 0.07737 | 0.32365 | 39077 | 12647 | 163464 | 359152 | 9.19 | 2.48 |
| 80 | 0.10644 | 0.41739 | 26430 | 11031 | 103638 | 195688 | 7.40 | 2.42 |
| 85 | 0.16728 | ... | 15398 | 15398 | 92050 | 92050 | 5.98 | 5.98 |
| Female |  |  |  |  |  |  |  |  |
| 0 | 0.03365 | 0.03271 | 100000 | 3271 | 97214 | 7125298 | 71.25 | 0.15 |
| 1 | 0.00072 | 0.00287 | 96729 | 278 | 386214 | 7028084 | 72.66 | 1.47 |
| 5 | 0.00045 | 0.00226 | 96451 | 218 | 481712 | 6641870 | 68.86 | 2.50 |
| 10 | 0.00033 | 0.00164 | 96233 | 158 | 480772 | 6160158 | 64.01 | 2.50 |
| 15 | 0.00094 | 0.00469 | 96076 | 451 | 479361 | 5679386 | 59.11 | 2.74 |
| 20 | 0.00106 | 0.00530 | 95625 | 507 | 476878 | 5200025 | 54.38 | 2.54 |
| 25 | 0.00115 | 0.00574 | 95118 | 546 | 474264 | 4723147 | 49.66 | 2.57 |
| 30 | 0.00150 | 0.00747 | 94572 | 706 | 471179 | 4248883 | 44.93 | 2.62 |
| 35 | 0.00208 | 0.01033 | 93866 | 970 | 467080 | 3777704 | 40.25 | 2.68 |
| 40 | 0.00364 | 0.01807 | 92896 | 1679 | 460538 | 3310624 | 35.64 | 2.65 |
| 45 | 0.00445 | 0.02200 | 91217 | 2007 | 451307 | 2850086 | 31.24 | 2.62 |
| 50 | 0.00671 | 0.03304 | 89211 | 2948 | 439200 | 2398779 | 26.89 | 2.67 |
| 55 | 0.01101 | 0.05366 | 86263 | 4629 | 420573 | 1959579 | 22.72 | 2.68 |
| 60 | 0.01770 | 0.08499 | 81634 | 6938 | 391873 | 1539006 | 18.85 | 2.65 |
| 65 | 0.02711 | 0.12738 | 74696 | 9515 | 350998 | 1147133 | 15.36 | 2.64 |
| 70 | 0.04482 | 0.20235 | 65181 | 13189 | 294272 | 796135 | 12.21 | 2.60 |
| 75 | 0.06908 | 0.29489 | 51992 | 15332 | 221929 | 501863 | 9.65 | 2.52 |
| 80 | 0.09822 | 0.39301 | 36660 | 14408 | 146687 | 279933 | 7.64 | 2.46 |
| 85 | 0.16700 | ... | 22252 | 22252 | 133246 | 133246 | 5.99 | 5.99 |

